# An Unsupervised XAI Framework for Dementia Detection with Context Enrichment

**DOI:** 10.1101/2025.05.28.25327435

**Authors:** Devesh Singh, Yusuf Brima, Fedor Levin, Martin Becker, Bjarne Hiller, Andreas Hermann, Irene Villar-Munoz, Lukas Beichert, Alexander Bernhardt, Katharina Buerger, Michaela Butryn, Peter Dechent, Emrah Düzel, Michael Ewers, Klaus Fliessbach, Silka D. Freiesleben, Wenzel Glanz, Stefan Hetzer, Daniel Janowitz, Doreen Görß, Ingo Kilimann, Okka Kimmich, Christoph Laske, Johannes Levin, Andrea Lohse, Falk Luesebrink, Matthias Munk, Robert Perneczky, Oliver Peters, Lukas Preis, Josef Priller, Johannes Prudlo, Diana Prychynenko, Boris S. Rauchmann, Ayda Rostamzadeh, Nina Roy-Kluth, Klaus Scheffler, Anja Schneider, Louise Droste zu Senden, Björn H. Schott, Annika Spottke, Matthis Synofzik, Jens Wiltfang, Frank Jessen, Marc-André Weber, Stefan J. Teipel, Martin Dyrba, the ADNI, AIBL, FTLDNI study groups

**Affiliations:** German Center for Neurodegenerative Diseases (DZNE), Rostock/Greifswald, Germany; Institute for Visual and Analytic Computing, University of Rostock, Germany; Translational Neurodegeneration Section “Albrecht Kossel”, Department of Neurology, University Hospital Rostock, Rostock, Germany; German Center for Neurodegenerative Diseases (DZNE), Berlin, Germany; Charité – Universitätsmedizin Berlin, corporate member of Freie Universität Berlin and Humboldt Universität zu Berlin, Department of Psychiatry and Neuroscience, Hindenburgdamm 30, 12203 Berlin, Germany; Division Translational Genomics of Neurodegenerative Diseases, Hertie Institute for Clinical Brain Research and Center of Neurology, University of Tübingen, Tübingen, Germany; German Center for Neurodegenerative Diseases (DZNE), Munich, Germany; Department of Neurology, University Hospital of Munich, Ludwig-Maximilians-Universität (LMU) Munich, Munich, Germany; Institute for Stroke & Dementia Research, University Hospital, LMU Munich, Germany; German Center for Neurodegenerative Diseases (DZNE), Magdeburg, Germany; Institute for Cognitive Neurology and Dementia Research, Faculty of Medicine, University Hospital Magdeburg, Magdeburg, Germany; German Center for Neurodegenerative Diseases (DZNE), Bonn, Germany; Department for Neurodegenerative Diseases and Gerontopsychiatry, University of Bonn, Bonn, Germany; Department of Psychosomatic Medicine, Rostock University Medical Center, Rostock, Germany; German Center for Neurodegenerative Diseases (DZNE), Tübingen, Germany; Section for Dementia Research, Hertie Institute for Clinical Brain Research, Department of Psychiatry and Psychotherapy, University Hospital Tübingen, Tübingen, Germany; Munich Cluster for Systems Neurology (SyNergy), Munich, Germany; Department of Psychiatry and Psychotherapy, University Hospital Tübingen, Tübingen, Germany; Department of Psychiatry and Psychotherapy, University Hospital, LMU Munich, Munich, Germany; Ageing Epidemiology Research Unit, School of Public Health, Faculty of Medicine, Imperial College London, London, United Kingdom; Department of Psychiatry and Psychotherapy, School of Medicine and Health, Technical University of Munich, Germany; University of Edinburgh and UK Dementia Research Institute, Edinburgh, United Kingdom; Department of Neurology, University Medical Centre, Rostock, Germany; Sheffield Institute for Translational Neuroscience, The University of Sheffield, Sheffield, United Kingdom; Department of Neuroradiology, University Hospital, LMU Munich, Germany; Department of Psychiatry, University of Cologne, Medical Faculty, Cologne, Germany; German Center for Neurodegenerative Diseases (DZNE), Goettingen, Germany; Department of Psychiatry and Psychotherapy, University Medical Center Goettingen, Goettingen, Germany; Neurosciences and Signaling Group, Institute of Biomedicine (iBiMED), Department of Medical Sciences, University of Aveiro, Aveiro, Portugal; Cologne Excellence Cluster on Cellular Stress Responses in Aging-Associated Diseases, Faculty of Medicine, University of Cologne, Cologne, Germany; MR-Research in Neurosciences, Department of Cognitive Neurology, University Medical Center Goettingen, Goettingen, Germany; Berlin Center for Advanced Neuroimaging, Charité University Medicine Berlin, Berlin, Germany; Department of Neurology, University Hospital Bonn, Bonn, Germany; Department for Biomedical Magnetic Resonance, University of Tübingen, Tübingen, Germany; Department of Psychiatry and Psychotherapy, University Hospital Magdeburg, Magdeburg, Germany; Institute of Diagnostic and Interventional Radiology, Pediatric Radiology and Neuroradiology, University Medical Centre Rostock, Rostock, Germany; Charité – Universitätsmedizin Berlin, corporate member of Freie Universität Berlin and Humboldt Universität zu Berlin, Experimental and Clinical Research Center (ECRC), Lindenberger Weg 80, 13125 Berlin, Germany; German Center for Mental Health (DZPG), Munich, Germany; Department of Psychiatry and Psychotherapy, Charité – University Medicine Berlin, Berlin, Germany

**Keywords:** Neurodegenerative diseases, Alzheimer’s disease, frontotemporal dementia, magnetic resonance imaging, brain volumetry, explainable artificial intelligence (XAI), qualitative evaluation

## Abstract

**Introduction:** Explainable Artificial Intelligence (XAI) methods enhance the diagnostic efficiency of clinical decision support systems by making the predictions of a convolutional neural network’s (CNN) on brain imaging more transparent and trustworthy. However, their clinical adoption is limited due to limited validation of the explanation quality. Our study introduces a framework that evaluates XAI methods by integrating neuroanatomical morphological features with CNN-generated relevance maps for disease classification.

**Methods:** We trained a CNN using brain MRI scans from six cohorts: ADNI, AIBL, DELCODE, DESCRIBE, EDSD, and NIFD (N=3253), including participants that were cognitively normal, with amnestic mild cognitive impairment, dementia due to Alzheimer’s disease and frontotemporal dementia. Clustering analysis benchmarked different explanation space configurations by using morphological features as proxy-ground truth. We implemented three post-hoc explanations methods: i) by simplifying model decisions, ii) explanation-by-example, and iii) textual explanations. A qualitative evaluation by clinicians (N=6) was performed to assess their clinical validity.

**Results:** Clustering performance improved in morphology enriched explanation spaces, improving both homogeneity and completeness of the clusters. Post hoc explanations by model simplification largely delineated converters and stable participants, while explanation-by-example presented possible cognition trajectories. Textual explanations gave rule-based summarization of pathological findings. Clinicians’ qualitative evaluation highlighted challenges and opportunities of XAI for different clinical applications.

**Conclusion:** Our study refines XAI explanation spaces and applies various approaches for generating explanations. Within the context of AI-based decision support system in dementia research we found the explanations methods to be promising towards enhancing diagnostic efficiency, backed up by the clinical assessments.

## Introduction

Alzheimer’s disease (AD) is a significant and growing burden on global healthcare systems. Estimates suggest a global population of 152.8 million people living with dementia by 2050 ^1^, for which AD accounts for more than two-thirds of all cases ^2^. The increasing prevalence of AD warrants the development of automated clinical decision support systems to improve the efficiency of diagnostic procedures and early disease detection. Deep learning (DL) has emerged as a promising tool in this context, offering state-of-the-art methods for the fast and robust analysis of complex neuroimaging data. However, the integration of DL into clinical practice is often hampered by a lack of transparency and interpretability of its predictions due to its ’black-box’ nature^3^.

Explainable Artificial Intelligence (XAI) methods offer a potential solution to this challenge by making DL models more human-comprehensible and interpretable. By explaining the decisions made by complex DL systems, XAI aims to bridge the gap between model predictions and clinical insights. This is particularly relevant under the European Union’s General Data Protection Regulation (GDPR) and Artificial Intelligence (AI) act, which under the ’right to explanation’ requires AI systems to provide explanations of their decision-making processes^4^. Other regulatory and government bodies have also advocated for similar AI capabilities, emphasizing the need for accountable and transparent AI systems in critical domains such as healthcare and medical decision-making^5–9^.

Despite the advancements in XAI methods, a research gap remains in validating and assessing the quality of the explanations generated by AI systems^10–12^. Notably, it is time-consuming and expensive to consult experts to provide ‘ground-truth’ explanations and to evaluate the explanations generated by XAI methods. It also requires additional fine-tuning of the XAI methods to improve their correctness and suitability for a specific use case. Furthermore, with regards to the inference process, it is often unclear and depends on the user’s experience level in determining—what needs to be explained, how, and in what detail^12^.

Additional methods for generating explanations rely on sets of rules, to combine symbolic reasoning such as knowledge graphs, with neural models to provide human-understandable insights into AI decision-making. Rule-based explanations offer structured and semantic explanations which enhances transparency by relying upon a-priori domain knowledge ^13,14^. Meanwhile, XAI methods that simplify model predictions reduce cognitive burden on the user by presenting the most useful information, and utilize methods such as network pruning or compression^15,16^. XAI methods like explanations-by-example describe model decision for a query sample by providing information about the most similar sample(s) from the training set ^17^.

Moreover, XAI-generated explanations, beyond the end use-case of providing insights into the AI system, may also be used as an additional information modality. One recent study highlighted a more separable arrangement of the participants in the ‘explanation space’, i.e., the vector space where the data points are arranged according to the explanation, compared to the original input space ^18^. Specifically for convolutional neural networks (CNN) applications, explanation space refers to the representational space derived from the attribution (relevance) heatmaps or its feature-level representations. Some CNN studies further explored this ‘quantification gap’ of explanations and evaluated the overlap between visual explanations, i.e., attribution-based relevance maps and ground truth ^19–21^. Other studies have addressed the consistency and coherence of the relevance mapping techniques with respect to expert-created ground truth segmentations ^22^. Some dementia studies quantified the voxel-level overlap between relevance maps and proxy ground truth maps, i.e., AD likelihood maps with relevant regions found through literature meta-analysis ^23,24^. Notably, all of these studies have predominantly used supervised machine learning approaches that utilize expert-assessed, expensive-to-obtain, ground truth or other proxy measures to calculate these ground truths.

Here, we propose to extend the common understanding of the explanation space and present a framework that incorporates clinically relevant morphological features - such as cortical thickness and gray matter volumetry - combined with relevance maps to create a *context-enriched explanation space.* Previous multimodal studies have informed our additive approach to extend the explanation space. To date, studies of dementia detection that combine multiple data modalities, such as MRI and PET scans, often outperform unimodal models ^25,26^. Previously developed disease state indices for differential diagnosis also utilized different information sources in a generalized additive model ^27^. Taken together, we hypothesized that the explanation space that better separates, distributes, and structures disease pathology information would also be a more appropriate space for generating explanations. We assumed that the combination of these information sources would produce more contextually sensitive explanations, which in turn would improve the quality of the explanations. To examine these assumptions, we performed a clustering analysis to explore the distribution of participants in different explanation space configurations. Our unsupervised analysis was intended to act as a proxy measure of the utility of explanations, thus bypassing the dependence of a supervised analysis based on ground truth explanation labels. Our framework generates *post hoc* explanations for a CNN model detecting dementia diseases at both global level, i.e., subgroup membership, and local level, i.e., cognitive trajectory examples or textual prediction explanations.

To comprehensively evaluate our AI-based explanations, we also conducted a qualitative analysis with expert clinicians, assessing each explanation’s usefulness for improving patient examinations. Through expert evaluation, we tackled a common issue in developing XAI prototypes for clinical decision support systems, i.e., the lack of user involvement in the co-development process^11,24,28^. Our overall aim was to advance the development and validation of robust XAI methods, and address the gaps in the evaluation of the explanations generated in the context of AD diagnosis.

## Methods

The workflow of our study is schematically presented in Figure 1. Our framework provides several ways to generate post-hoc explanations for a CNN model trained to detect dementia diseases, including: i) global-level explanations, such as membership in the stable versus converter subgroups, and ii) local-level explanations for each individual prediction, such as ii-a) example-based explanations of cognitive trajectories or ii-b) textual explanation by pathology summarization. To evaluate clinical validity of the different types of AI-based explanations produced from our framework, we also conducted a qualitative analysis with a focus group of radiologists (N=4) and neurologists (N=2).

**Figure 1:**
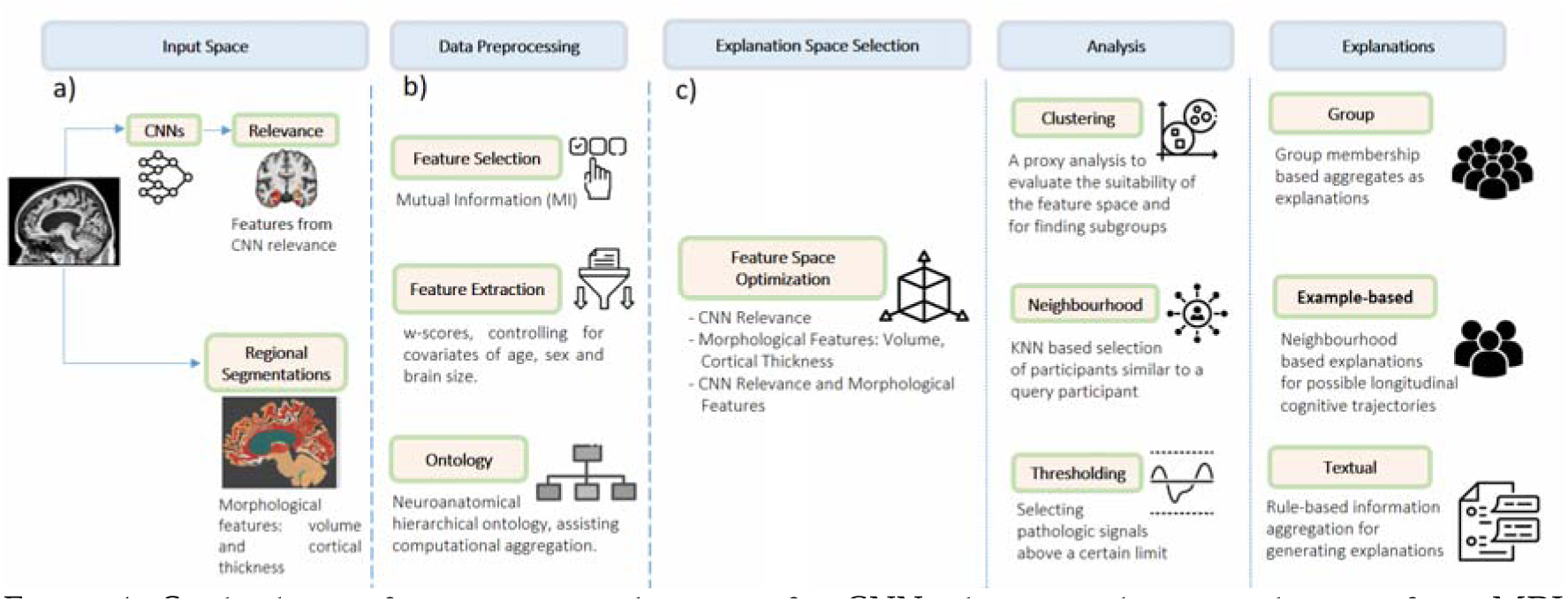
Study design for creating explanations for CNNs detecting dementia diseases from MRI scans. Here we illustrate a) the input space with trained CNN’s relevance maps and brain segmentation, b) the preprocessing steps of - feature selection and extraction, and c) the explanation generation from the context-enriched explanation space and features extracted, utilizing different analysis methods.

### Neuroimaging datasets

In this study, we collected T1-weighted brain MRI scans (N=3253) from publicly available neuroimaging data cohorts. The data scans were pooled from the following data cohorts: i) the Alzheimer’s Disease Neuroimaging Initiative (ADNI), study phases ADNI2/GO and ADNI3, ii) the Australian Imaging, Biomarker & Lifestyle Flagship Study of Ageing (AIBL) ^29^, iii) the DZNE Longitudinal Study on Cognitive Impairment and Dementia (DELCODE) ^30^, iv) the European DTI Study on Dementia (EDSD) ^31^, v) the DZNE Clinical Registry Study on Frontotemporal Dementia (DESCRIBE-FTD), and vi) the Frontotemporal Lobar Degeneration Neuroimaging Initiative (FTLDNI) which is also known as Neuroimaging Initiative in Frontotemporal Dementia (NIFD).

It should be noted that mild cognitive impairment (MCI) can arise from various underlying conditions, however, the ADNI, AIBL, and DELCODE cohorts apply inclusion and exclusion criteria to focus primarily on amnestic MCI, i.e., individuals with memory impairment. Other conditions, such as depression or substance abuse, were excluded. Summary statistics for the data used are presented in Table 1. See Supplementary Table A.1.1 for statistics reported for each cohort.

**TABLE 1.** Sample Statistics Per Disease Diagnosis Stage and Subtype.

|  | CN | MCI | AD | FTD |
| --- | --- | --- | --- | --- |
| **Sex (M/F)** | 732/963 | 448/369 | 266/282 | 112/81 |
| **Age** | 70.15 ± 7.58 | 72.76 ± 7.38 | 74.14 ± 7.69 | 63.35 ± 7.92 |
| **MMSE** | 29.02 ± 1.52 | 27.36 ± 2.3 | 22.15 ± 4.18 | 23.56 ± 7.10 |

CN: cognitively normal, MCI: mild cognitive impairment, AD: dementia due to Alzheimer’s disease (the typical presentation), FTD: Frontotemporal dementia where phenotypes include behavioral variant, semantic variant, and progressive nonfluent aphasia, MMSE: mini-mental state examination score, F: female, M: male. Numbers are reported as (mean ± s.d.).

These datasets were initially intensity corrected using the N4ITK algorithm for bias field correction. Then, HD-BET was applied for skull stripping^32^. ANTs SyNQuick registration tool was used to linearly warp all images to the MNI reference spare and ANTs AtroposN4 was applied for tissue segmentation into CSF, white matter, and gray matter. The normalized gray matter maps served as input for the CNN model. Subsequently, based on the native space images, FastSurfer version 2.0.4 was used to perform brain segmentation into 100 anatomically defined regions-of-interest (ROIs) and cortical surface reconstructions to measure regional volume and average cortical thickness^33,34^. FastSurfer follows the Desikan– Killiany–Tourville (DKT) atlas protocol for producing the anatomical segments^35,36^. Finally, the linear deformations from ANTs were applied to the FastSurfer segmentation maps to extract CNN relevance scores per region.

### Relevance segmentation, aggregation, and abstraction

We trained a multi-class CNN model based on the DenseNet architecture as the backbone ^37,38^. A three-way classification setup was used that classified cognitively normal (CN), Alzheimer’s disease (AD; pooled patients with dementia due to AD and patients with amnestic mild cognitive impairment (MCI)), and phenotypes of frontotemporal dementia (FTD) - including behavioral variant (bvFTD), semantic dementia (SD), and progressive nonfluent aphasia (PNFA) - participants. See supplementary section A.2 for further model training details.

We used the Layer-wise Relevance Propagation (LRP) attribution method that generates a heatmap of input regions that the model found useful for differentiating each class ^39^. We chose the composite alpha-beta LRP rule as it highlights relevant input features with high specificity and avoids the dispersed distribution of relevance scores across multiple input regions ^40–43^, unlike other methods such as GradCam ^44^ or Occlusion Maps ^45^. While the LRP rule is sensitive to the parametric choice of alpha and beta hyper-parameters, it does not require defining a base image used by methods like the Integrated Gradients ^43,46^. LRP relevance maps have also been used in several other previous dementia studies ^47–49^.

The 3D LRP relevance maps were segmented using region of interest (ROI) segmentations generated by the FastSurfer segmentation tool. Within each ROI, we calculated the relevance density, i.e., the relevances were summed up and divided by the volume of the respective ROI. The relevance density metric has previously been found to be better associated with disease features than the total sum or other relevance aggregation mechanisms ^50,51^. Based on the hierarchical ontology structure developed in our previous work^52^, relevance was aggregated and summarized across different levels of neuroanatomical abstraction, such as lobes and hemispheres; which added 24 higher-order (parent-level) aggregation concepts.

### Data preprocessing

#### Feature extraction

W-scores were calculated for each pathology feature, region, and participant, which quantified the relative deviations from the normative expectations. W-scores are an extension of Z-scores that adjust for covariates; in our study, we controlled for age, sex, brain size, and magnetic field strength, as these variables are widely known to influence brain volume and cortical thickness measures ^53–55^.

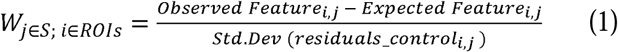

here *features*, i.e., S = {CNN relevance, cortical thickness, volume}. The expected feature is the prediction from a linear regression model that accounts for the confounding covariates and was trained only on the cognitively normal control participants. The residuals_controls_i,j_ are the residuals from the cognitively normal controls.

#### Feature selection

W-score features per region (X), i.e., the CNN’s relevances, the volumetric measure, and the cortical thickness measure, were compared with the disease diagnosis labels (Y), by calculating the mutual information I(X, Y) between them, defined as:

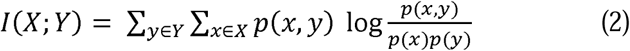

where in equation 2, p(X, Y) is the joint probability distribution for random variables X and Y, while p(X) is the marginal probability distribution for the random variable X. Mutual information quantifies the dependence between individual features and the disease diagnosis, and lies between [0, +∞) where a mutual information of 0 indicates two independent variables. Mutual information (MI) is one of the most widely used methods for feature selection in machine learning, as it effectively quantifies the dependency between features and target variables. Its ability to capture both linear and non-linear relationships makes it particularly valuable in high-dimensional data analysis ^56^. The features with mutual information above the threshold of 0.1, chosen heuristically, were selected for further analysis.

### Enriched explanation space

We set up a clustering analysis as a proxy measure to calibrate the suitability of the various explanation spaces. Different variations of the explanation space were explored: a) including only the relevance features from the CNN, i.e., the basic *explanation spac*e, b) including only the morphological features - volumetry and/or cortical thickness, and c) including both the relevance features from the CNN and the morphological features, i.e., the *context-enriched explanation space*. The derived clusters were evaluated using broadly two sets of metrics. First, the external validation metrics, measuring agreement between the predicted cluster labels and the ground truth disease diagnosis - homogeneity, completeness and v-measure ^57^, adjusted mutual information ^58^, adjusted rand score ^59,60^, and Fowlkes score ^61^. V-measure is the harmonic mean of the homogeneity and completeness scores. Second, the internal validation metrics, measuring the separation between the clusters within the space and requiring no external ground truth labels - average silhouette coefficient ^62^ and Davies Bouldin score ^63^.

### Deriving Explanations from the Enriched Explanation Space

#### Group-level explanations

We utilized the agglomerative hierarchical clustering with Ward’s linkage to create the group-level, ***feature simplification*** explanations. The hierarchical clustering separates different subgroups of participants. Ward linkage criterion minimizes the within-cluster variance ^64^ and has been found useful in other dementia studies ^65,66^. We chose the Euclidean distance as the metric for calculating the distance between the clusters in the explanation space. For the participants grouped within a cluster, a repeated-measures linear mixed-effect model was fitted to cognition trajectories for Mini-Mental State Examination (MMSE) and global Clinical Dementia Rating (CDR) scores. The models included fixed effects for age at baseline, sex, and interaction terms between baseline cognitive diagnosis (cognitively normal - CN, mild cognitive impairment - MCI, or Alzheimer’s disease dementia - AD), cluster index, and time (months). We also specified random intercepts for each participant to account for individual variability in baseline cognition. We additionally performed the Kaplan-Meier survival analysis to compare the time to dementia conversion between the clusters. The conversion event was marked by the change of the CDR global score, i.e., conversion from unimpaired cognition (CDR=0) to MCI due to AD (CDR= 0.5), and conversion from MCI (CDR=0.5) to mild AD dementia (CDR=1), beyond which any further increase in CDR score (>1) was not considered. For each participant, longitudinal data was included for up to six years, and participants were right-censored when they did not convert.

#### Example-based explanations

The example-based explanations were generated using a meta-classifier abstracting over the details of the explanation space and presenting the likely cognition progression trajectories. The use of a simple meta-classifier is a common practice in building decision support systems to assist experts ^67^. We chose the k nearest neighbor (KNN) classifier as the meta-classifier. The size of the neighborhood k=10 was set heuristically. The nearest neighbor for a query sample in the enriched explanation space represents a small group of examples, i.e., participants with similar pathology. Hence, using this notion of similarity, we then present exemplary cognition trajectories of the neighbors, here, the MMSE and CDR scores obtained from follow-up visits up to six years, where available.

#### Textual explanations

Previous studies that applied knowledge-based approaches ^68–70^ have established a more structured and knowledge-engineered usage of clinical information for decision support. We previously created a computational neuroanatomy ontology that enhanced the aggregation of pathologic information^52^. Based on this framework, we developed a post-hoc, ***rule-based*** explanation method where the information sources, here CNN relevances, volume, and cortical thickness features, could be integrated to generate ***textual explanations*** for a single participant.

The ontology’s hierarchical structure opens up space for the computational aggregation of different pathological features, at multiple abstraction levels. More importantly, the structured setup also allows for more sophisticated logical reasoning, for example, the inclusion or exclusion of entities. We developed a rule-based method that dynamically chooses anatomical entities for which all three (logical and) pathological features indicated abnormal levels, more specifically, the w-score exceeding 2 standard deviations from the norm. In cases where many regions at a lower hierarchy were selected, then only the higher hierarchy region was selected for presentation. This reduces the load of information presented to the end user.

The selected regions were reported to the clinical users as template-based textual explanations. When the average pathology w-score across all applicable features remains between 2 and 3, a region is classified as ‘mild’ pathology. Scores between 3 and 4 indicate ‘moderate’ pathology, while, scores exceeding 4 standard deviations indicate ‘strong’ pathology. This threshold-based logic was empirically derived through the analysis of w-scores in our dataset. This logic facilitates the categorization of average pathology severity for each neuroanatomic region.

### Qualitative interviews with the experts

We interviewed neurologists (N=2) working in a memory clinic and radiologists (N=4) with an average of 10+ years of experience. This ensured expert feedback while avoiding input from newly trained professionals. The semi-structured interview was opened with introducing the experts to the high-level perspectives present in explainable artificial intelligence (XAI) field, i.e., the different values and goals, such as causality, confidence, informativeness, and trustworthiness, pursued by the XAI methods ^71^. They were also introduced to the taxonomies for grouping various XAI methods ^16^.

Furthermore, the semi-structured interview consisted of the following steps: introducing a case study sample, the various types of explanations generated for the sample presented one by one, and prompting the experts regarding the different usability aspects of the explanations. Figure 2 illustrates the process of semi-structured expert interviews. All interviews were carried out in accordance with relevant guidelines and regulations. For further details, please refer to the included ethics statement.

**Figure 2.**
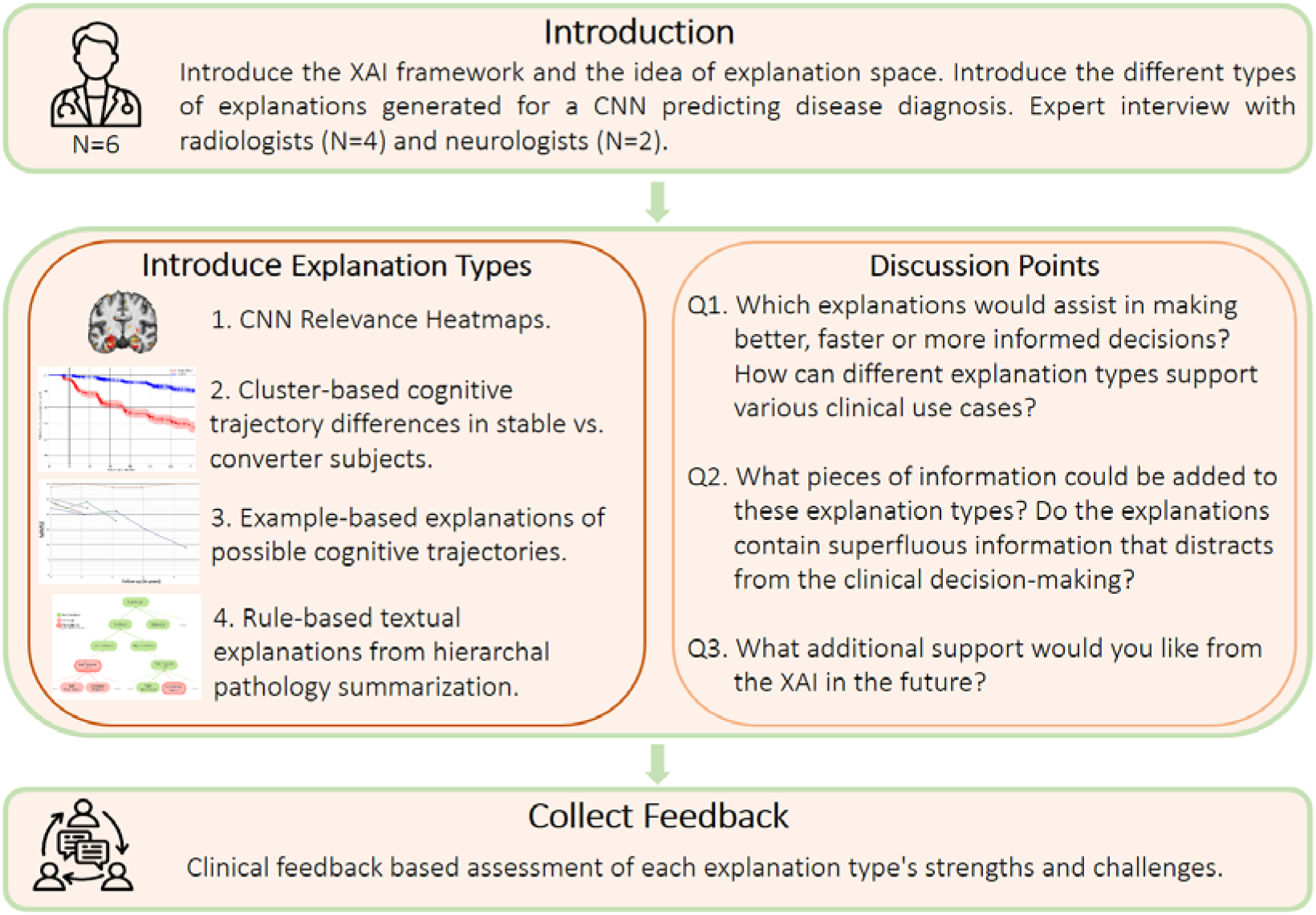
Semi-structured expert interview flowchart. We collected the clinical feedback on different aspects of XAI explanation types to assist clinical decision-making. The interviews approximately lasted for an hour. Clinical experts highlighted which explanations enhance the decision-making process by making CNN more adoptable, what information should be added or removed, and future improvements for XAI support. These interviews served as a basis for qualitatively evaluating the opportunities and challenges of applying XAI methods.

This order was chosen to let the experts present their opinions about each method’s value for a use-case, building on the opinions to define the more concrete strengths and challenges, and eventually to state the possible future works for the explanation types. Approximately an hour to an hour and a half was spent to go through all the explanation types and discuss them individually. The focus group interviews were conducted one-on-one or with at most two experts together. In total, 4 interview sessions were conducted. The interviews were conducted between February and March 2025. The focus group interviews helped us qualitatively evaluate the opportunities and challenges of applying XAI methods in clinical decision systems.

## Results

### Explanation Space Selection and Subcluster Identification

From our CNN model trained for three-way classification between CN, AD, and FTD, the CN node was chosen to acquire the relevance, as the relevance scores generated from it represent the deviation from the normal group. This means that the relevance of an input voxel reflects its contribution to a subject being classified (or not classified) as cognitively normal, thereby highlighting patterns associated with pathological aging.

After applying a heuristically set threshold of 0.1 on the mutual information criterion, 81 features remained. The selected features included K=19 (23.5%) relevance features, K=46 (56.7%) volume features, and K=16 (19.8%) cortical thickness features. Notably, for the cortical regions left entorhinal, left inferior and superior temporal, and left temporal lobe, as well as for the subcortical regions left hippocampus, left putamen, and left amygdala, all respective features had mutual information above the threshold. See Supplementary section A.3 for more information.

Using the selected features, agglomerative clustering with ward linkage was conducted to compare different variations of the explanation space while separating the AD and CN participants. The results of the clustering analysis are presented in Table 2. According to the V-measure, the enriched explanation space provided the highest score of 0.43.

**TABLE 2.**
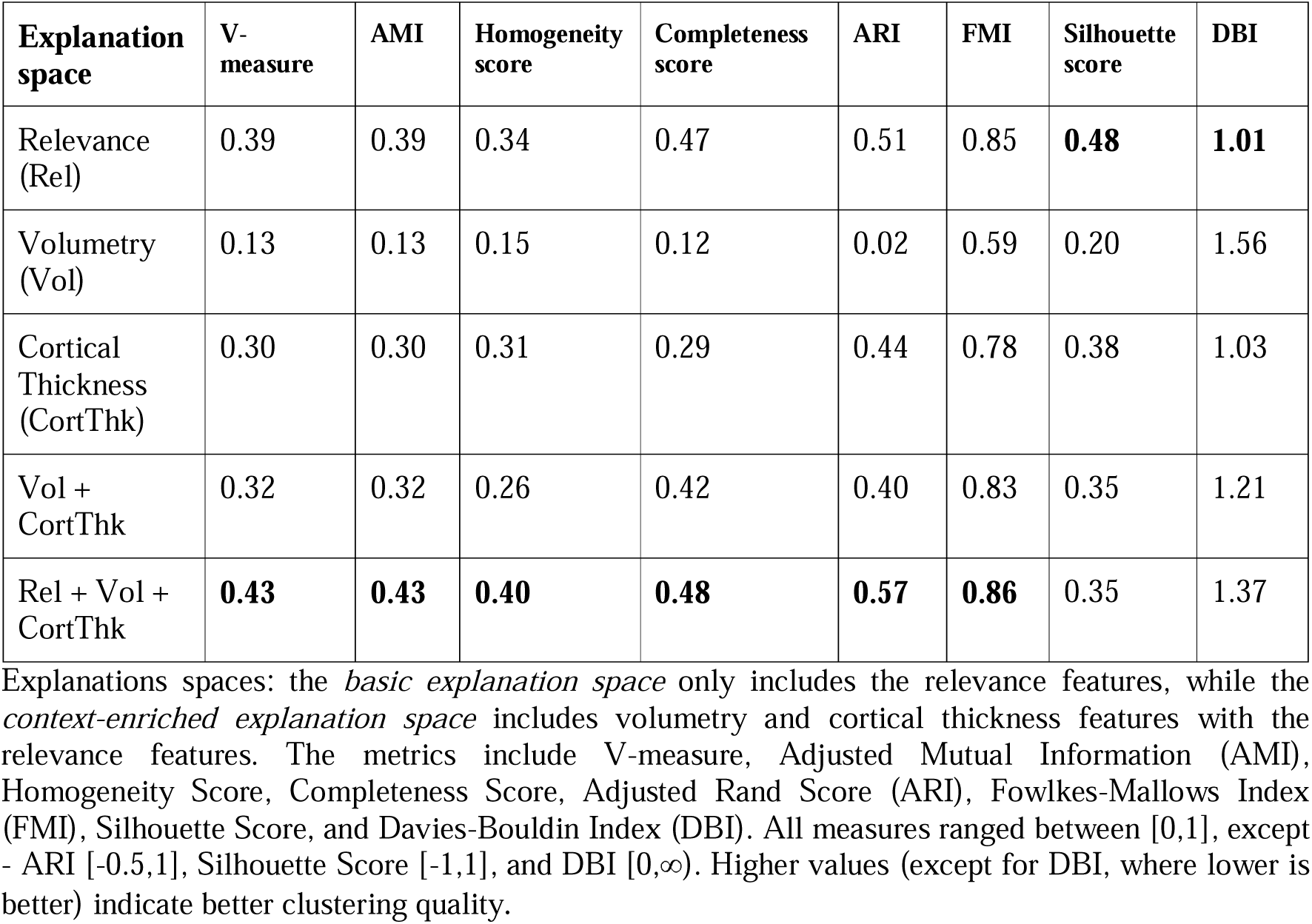
Clustering Performance Across Explanation Spaces.

Figure 3 illustrates the cluster map of the dataset in the context-enriched explanation space, providing a hierarchical visualization (on the Y-axis) of relationships between data points. From the heatmap intensity, we found a relative segregation of the disease diagnoses between the two main clusters, where darker regions on the heatmap represent more pathologic patients being clustered together. The number of clusters was heuristically set to two to balance between cohesion and separation while maintaining clinical interpretability. Although we explored 3-4 clusters scenarios based on the splits found via the dendrogram, they did not provide additional meaningful insights. While a relatively homogeneous cluster with FTD patients emerged (in 3 cluster scenario), it offered limited new information with respect to the further explanations drawn from the framework.

**Figure 3.**
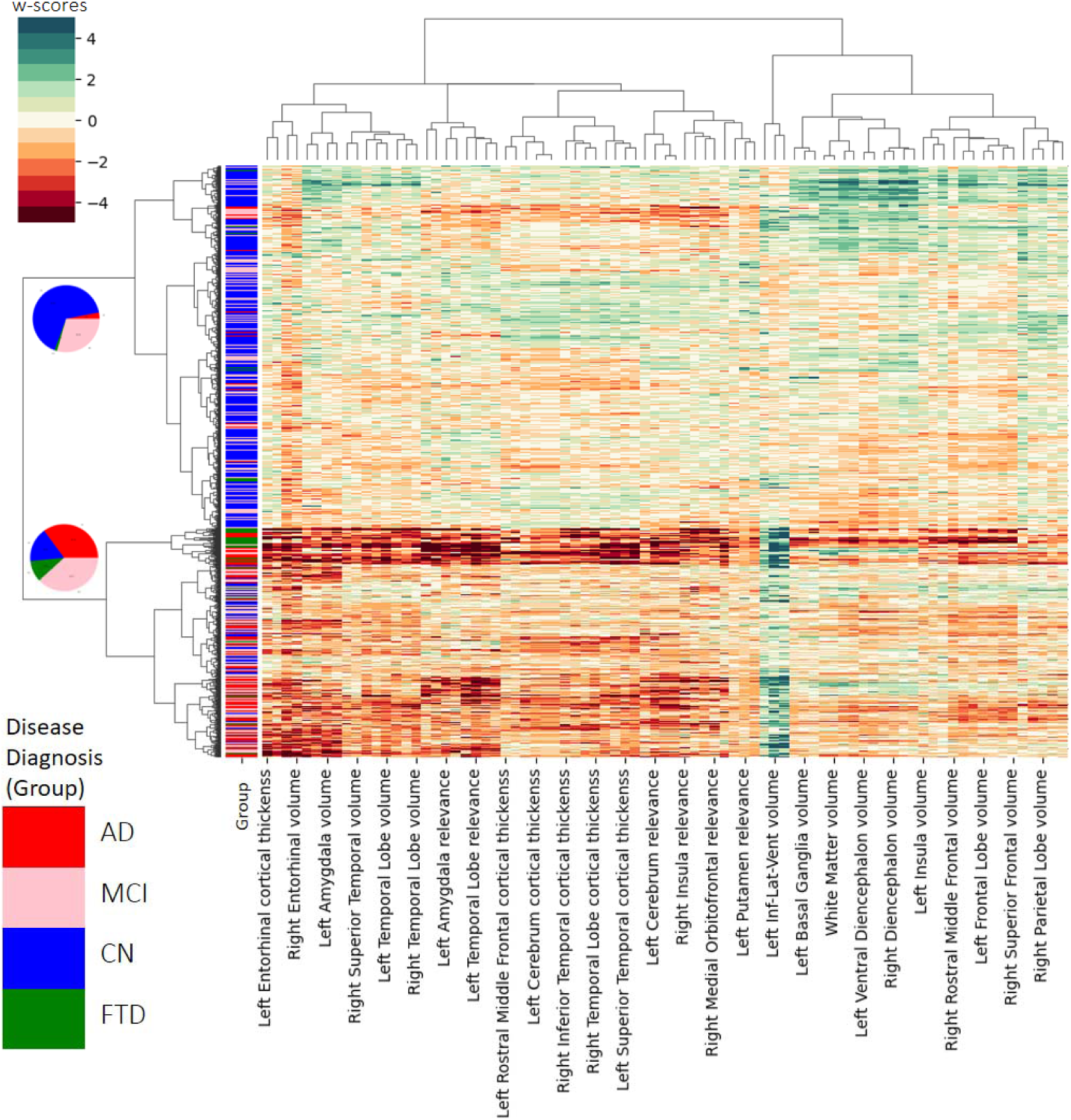
Cluster Map. Hierarchical clustering dendrogram (on the Y-axis) resulting from Ward’s hierarchical clustering analysis of individual w-scores profiles of participants computed from three features - CNN relevances, volume, and cortical thickness measures. Four disease diagnoses were considered: cognitively normal (CN, color-coded as blue), mild cognitive impairment (MCI, color- coded as pink), dementia due to Alzheimer’s disease (AD, color-coded as red), and frontotemporal dementia (FTD, color-coded as green). The pie charts visualize the relative homogeneity, with respect to the disease diagnoses, of the two clusters. The W-score features are visualized using a custom color scale to indicate the extent of the deviation, where a gradual intensification of color (either red or green) signifies increasing pathological observations. For a vector graphic rendering, please refer to the GitHub version of the plot.

As visualized by the pie charts in the left dendrogram in Figure 3, one cluster mainly consists of healthy controls or participants with low levels of pathology, this cluster is here on termed as the *stable* cluster. The second cluster consists of participants with more advanced pathology, i.e, a high amount of atrophy in dementia patients, this cluster is here on termed as the *converter* cluster. Table 3 presents the confusion matrix comparing clustering outcomes with ground truth labels of participants’ baseline disease diagnosis.

**TABLE 3.** Confusion Matrix for the Clustering Outcome.

| **Clusters /**<br>**Disease Diagnosis** | **CN** | **MCI** | **AD** | **FTD** |
| --- | --- | --- | --- | --- |
| Stable Cluster<br>(N=2084) | 1383<br>(66.4%) | 612<br>(29.4%) | 63<br>(3%) | 26<br>(1.2%) |
| Converter Cluster<br>(N=1318) | 219<br>(16.6%) | 503<br>(38.2%) | 462<br>(35.1%) | 134<br>(10.2%) |

The proportion of the participant's baseline disease diagnosis stage and subtype within the found clusters - termed *stable* or *converter* - are presented as percentage points. CN: cognitively normal, MCI: mild cognitive impairment, AD: dementia due to Alzheimer's disease (the typical presentation), FTD: Frontotemporal dementia where phenotypes include behavioral variant, semantic variant, and progressive nonfluent aphasia.

Using Fleiss’ Kappa (κ), a score for inter-rater reliability was calculated to evaluate the stability (agreement) for the clustering-based binary classification task of stable vs. converter. We performed a 4-fold cross-validation, yielding a κ score 0.77, indicating substantial agreement, i.e., a relatively stable clustering outcome that is independent of the data folds used for initialization.

### Simplified, Group-Level Explanations

Based on the two clusters identified in the context-enriched explanation space, the longitudinal cognitive trajectories were explored as simplified explanations of the CNN model’s predictions. Analysis of the longitudinal MMSE scores showed that the two identified clusters separate participants which would remain relatively stable or decline at an accelerated rate, i.e., converters. Specifically, as seen in Figure 4, the MMSE score of the high-risk converter group exhibited cognitive decline at a rate of 0.54 points per year, whereas the low-risk stable group declined at a rate of 0.02 points per year. Details for the mixed-effects modeling and the analysis of the Clinical Dementia Rating (CDR) global score can be found in the supplementary section A.4.

**Figure 4:**
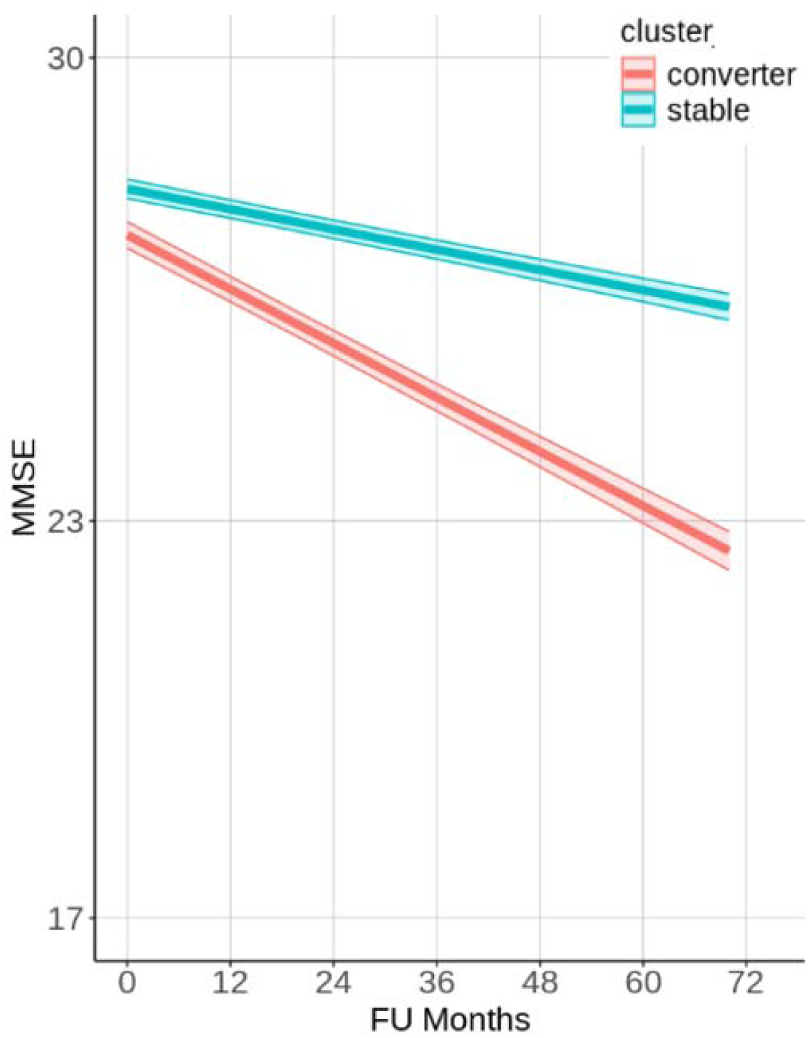
Longitudinal cognitive trajectories of different clusters of participants identified in the context-enriched explanation space. Values on MMSE cognitive test are obtained from mixed effects regression models which included the age, sex, baseline disease diagnosis, and the interaction between cluster membership and follow-up time in months (FU Months), as well as the interaction between baseline disease diagnosis and follow-up months. The model also included random intercepts for each participant to account for repeated measurements. The shaded regions represent 95% confidence intervals.

From the Kaplan-Meier survival analysis, we also see a similar separation (Figure 5), where 80% of the participants in the stable cluster remain free of conversion for 60 months (or 5 years), with a conversion rate of approximately 3.3% per year, while in the converter cluster approximately 10% of the participants convert per year.

**Figure 5:**
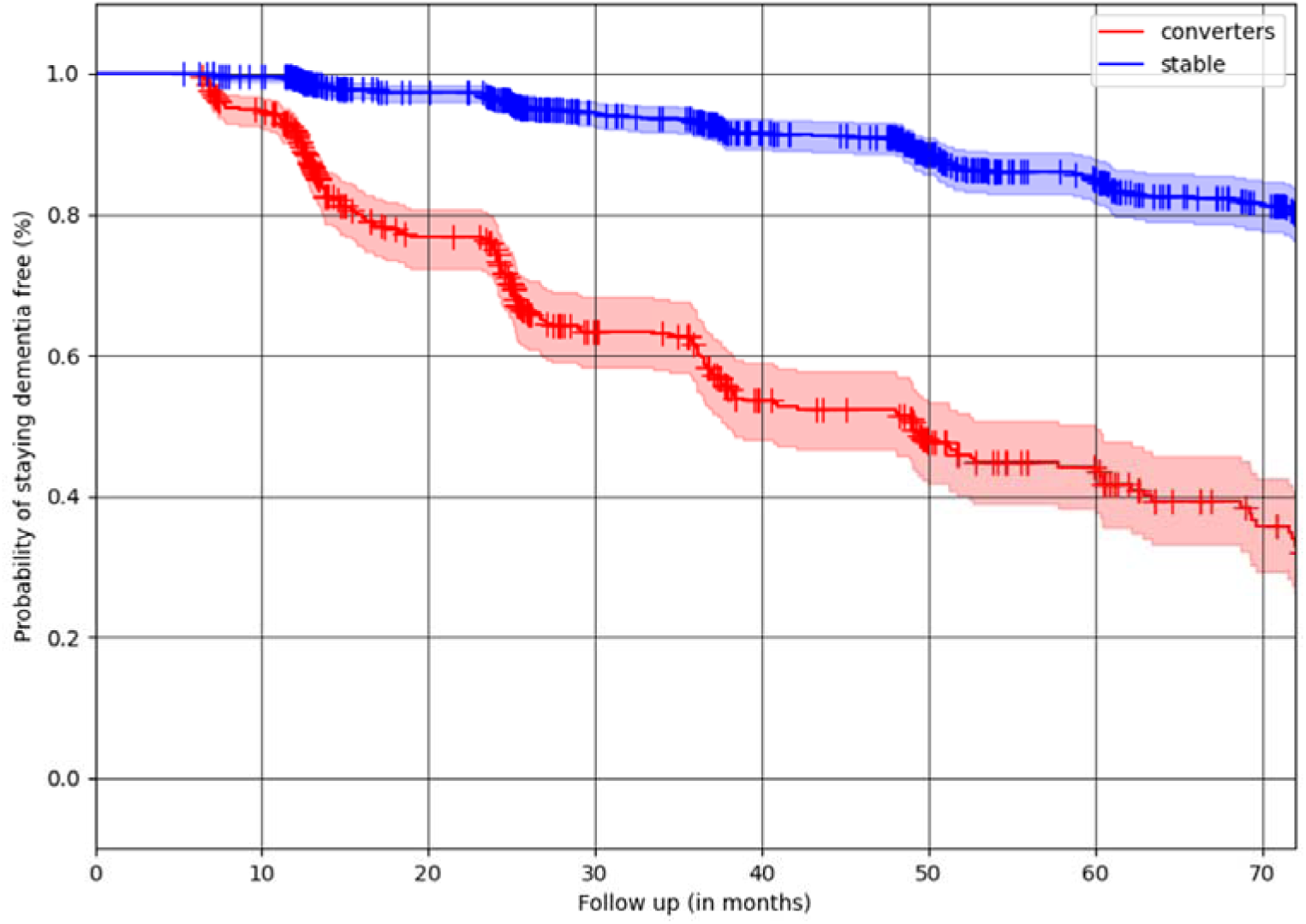
Kaplan-Meier curves illustrating the time to conversion across identified clusters. These survival curves represent the proportion of participants within each cluster who progressed either from CN to MCI and/or from MCI to dementia. Participants who did not develop dementia during the observation period were censored.

### Explanation by examples

Within the context-enriched explanation space, the longitudinal cognitive trajectories of participants with similar pathology to a query sample illustrate the possible trajectories over 72 months (6 years). A k nearest neighbor (KNN) model was employed, to find the participants that present the most similar pathology, with the neighborhood window heuristically set to k=10.

Figure 6 shows the cognitive trajectories on the MMSE cognitive test, based on the nearest neighbors of one arbitrarily selected individual from the DELCODE data cohort with the clinical diagnosis of MCI. Supplementary Figure A.5.1 illustrates the explanation-by-example cognitive trajectories for the CDR score, and Supplementary section A.6 illustrates MMSE and CDR explanation-by-example plots where each cognitive trajectory is shown in a unique color for more detail. The participant is a woman in her late sixties who has received almost ten years of formal education, and who has a baseline MMSE score of 24. In the figure legend, the ten nearest neighbors (with their baseline diagnosis and pseudonymised patient ID) are listed in the order of increasing Euclidian distance from this query sample, i.e., the most similar participant in the dataset is listed first. In a clinical setting, the trajectories could serve as illustrations of possible future cognitive development for the query participant.

**Figure 6:**
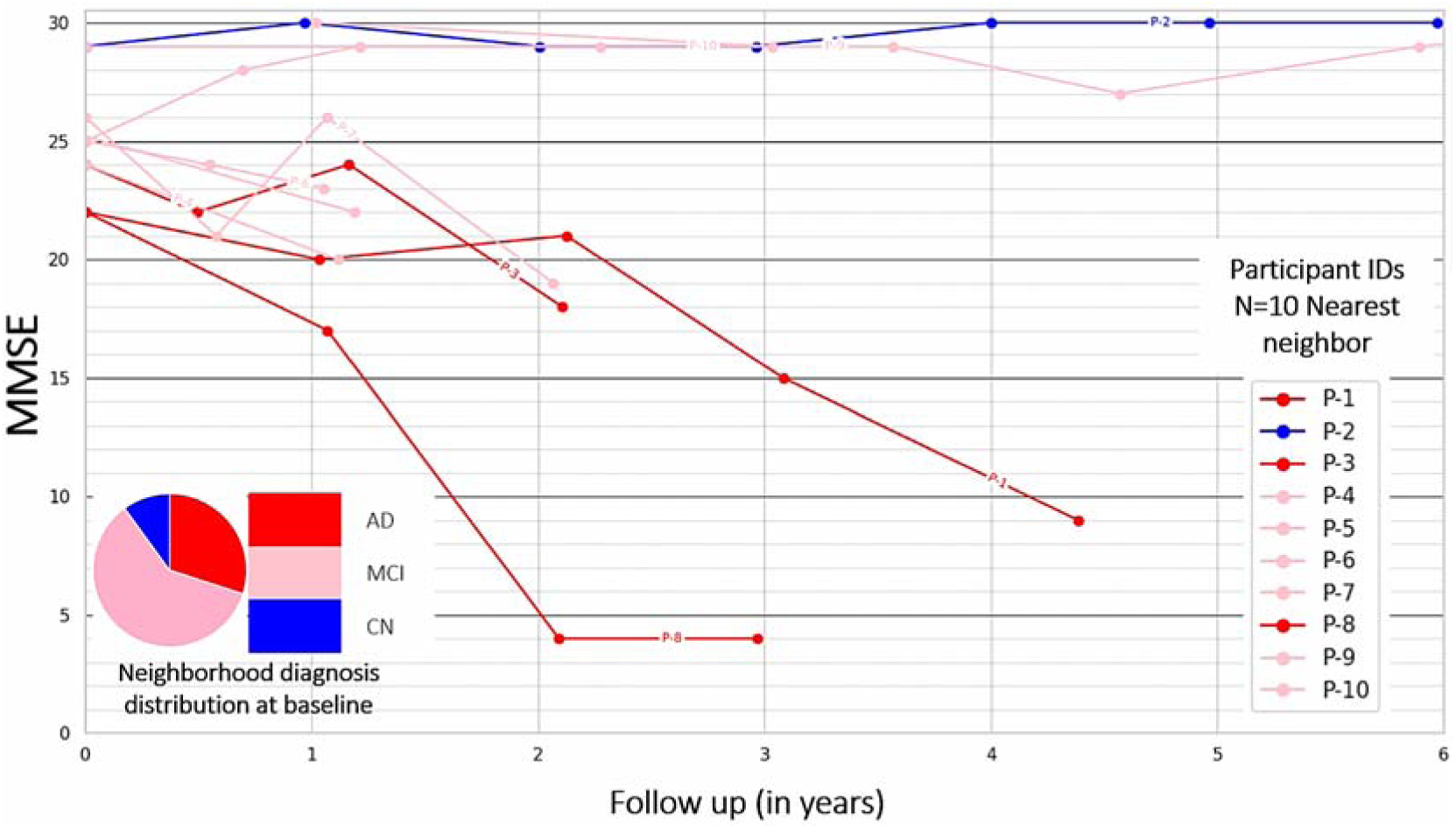
Explanation-by-examples: Within the context-enriched explanation space, the longitudinal cognitive trajectories of k=10 nearest neighbors of a query participant, from the DELCODE cohort, are shown. Scores on the MMSE cognitive test were observed on follow-up examinations for up to 6 years. Patient IDs of the nearest neighbors are pseudonymised, and the nearest neighbors are listed in the order of increasing Euclidian distance from the query sample, illustrating possible future cognition trajectories for the query participant. The cognition trajectories are additionally color-coded by the baseline disease diagnosis.

### Rule-based textual explanations

The knowledge-driven, ontology-based explanation method generated structured textual explanations for individual participants. By combining CNN relevances, volumetric, and cortical thickness measures, the rule-based mechanism generated hierarchical summaries of neuroanatomical abnormalities, reducing redundancy by prioritizing higher-order regions. In Figure 7 (a), we see an illustration of the hierarchical selection mechanism. Figure 7 (b,c) illustrates the template-based textual report generated for the same query participant from the DELCODE cohort. Figure 7 (b) lists all the pathologic regions identified, including the left superior temporal, left middle temporal, left temporal lobe, left inferior temporal, and left inferior lateral ventricle. Meanwhile in Figure 7 (c) generated a template-based summary, presenting pathologic information specifically for the left temporal lobe and left inferior lateral ventricle.

**Figure 7:**
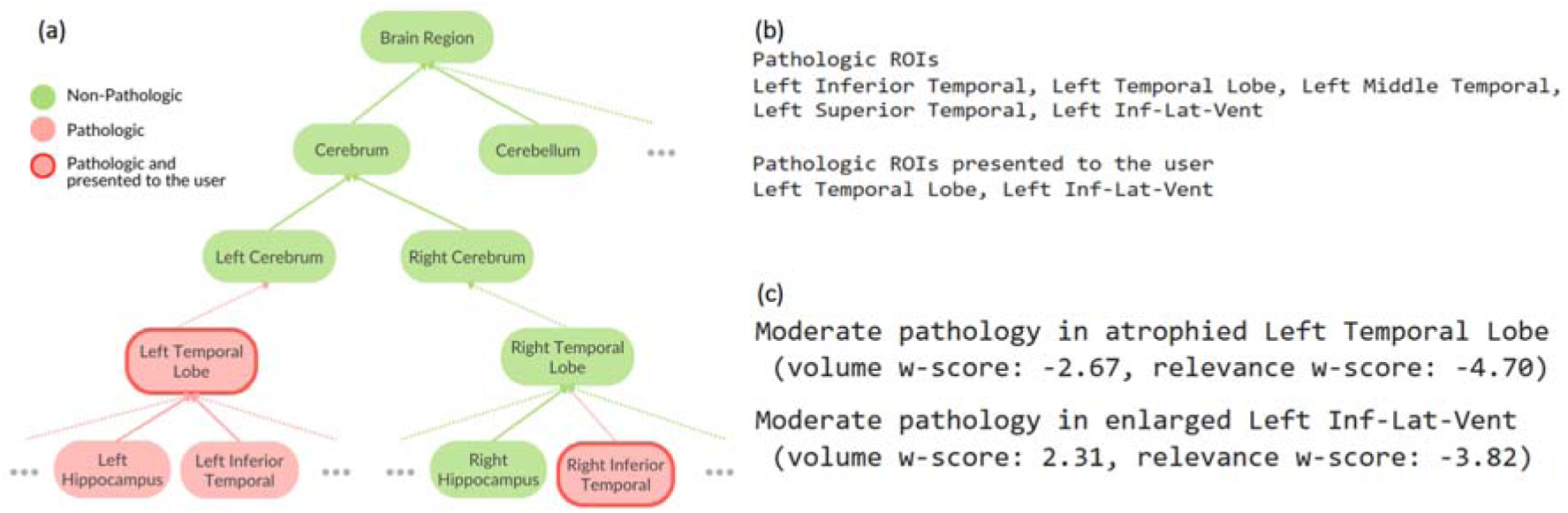
Rule-based textual explanation: (a) an hypothetical exemplary visual illustration of the rule-based mechanism of selecting neuroanatomical regions for which a pathologic threshold is reached for all the features - cortical thickness, volumetry, and relevance; and then narrowing down and optimizing the pathologic regions presented to the clinical user to reduce the information load. For a query participant from the DELCODE cohort, we show (b) a list of all the pathologic and presented regions, and (c) a template-based summary generated, listing w-scores from all relevant features.

### Qualitative evaluation of the explanation types

Neurologists (N=2) from the memory clinic found the simplified, group-level explanations to be particularly useful, as an aid to communicate with other clinical experts. They described a scenario where their risk assessment capabilities of XAI methods could possibly help in evaluating an individual’s eligibility for clinical trials. They also reported valuing the succinctness of these explanations, emphasizing the importance of limiting the presented information to 3–5 key facts to prevent cognitive overload. For patient interactions in memory clinics, explanation-by-example methods were seen as beneficial in facilitating personalized discussions, particularly to encourage healthier lifestyle choices such as quitting smoking, increasing social engagement, and exercising. However, neurologists also expressed reservations about using the explanation-by-example method with laypersons, as it could cause unnecessary anxiety to their patients, and acknowledged the inherent uncertainty in predicting an individual’s future cognitive development.

Radiologists (N=4) favored textual explanations, as these aligned well with their clinical workflow of reporting pathological findings across different regions of interest. They reported being in favor of XAI systems that could pre-identify relevant areas, potentially saving time by highlighting key regions before manual assessment. However, they found relevance heatmaps to be of limited utility, as these visualizations did not directly support their need for regional pathological descriptions. Radiologists in our study also requested that the XAI methods should align with disease diagnosis guidelines, e.g., from the German Society for Neurology (Deutsche Gesellschaft für Neurology), and should automatically highlight relevant brain regions based on the suspected pathology.

Beyond XAI method’s clinical validity, both radiologists and neurologists advocated for AI systems capable of integrating longitudinal patient data while accounting for comorbidities beyond neurodegeneration, such as depression, microbleeds, white matter lesions, and medical history, which may influence a patient’s current disease presentation. Neurologists highlighted the need for multi-disease diagnostic capabilities to assess the likelihood of different pathologies. Additionally, they expect XAI methods to quantify certainty and confidence intervals of their suggestions. Neurologists also requested an extension to the explanation-by-example approach, to incorporate multimodal data—including PET-Tau, blood-based biomarkers, and genetic makeup, to be more confident of the projected trajectories.

## Discussion

In this study, we introduced a framework that offers a novel unsupervised approach to XAI by extending the scope of conventional relevance heatmaps. Our study extends the basic explanation space by including the regional morphological information, i.e., cortical thickness and volumetry measures, creating the *context-enriched explanation space.* Within this new space, we quantified the information present in the relevance heatmaps and provided evidence for relatively better clustering outcomes with respect to the disease diagnosis labels. We also explored three different methods of generating explanations for the model’s predictions, namely: (i) group-based clustering of stable and converter participants, leading to simplified explanations, (ii) neighborhood-based examples of cognitive trajectories, and (iii) rule-based textual reports of pathologic regions. To the best of our knowledge, only a few studies have quantitatively compared relevance heatmaps between different dementia diagnosis groups^23,24,50^. Our study is the first of its kind to examine clinicians’ feedback for the generated explanation types.

While our framework offers an enriched feature space that integrates model-derived relevance maps with regional morphological measures, it does not directly capture the entire decision-making process of the CNN. Instead, it validates and contextualizes the information embedded in the relevance maps by situating them in a clinically interpretable feature space. Within the broader challenges of XAI in imaging, aside from certain preliminary approaches such as topographic activation maps^72,73^, no current method, to the best of our knowledge, is capable of exhaustively reconstructing or tracing the internal reasoning of CNNs or other deep models. Our contribution should therefore be seen as a complementary approach to the existing relevance heatmaps generating methods, in that it enhances interpretability by bridging abstract heatmaps with clinically meaningful features, while acknowledging the limitations in tracing deep models’ reasoning.

### Enriching explanation space and explanations generation

Recent studies have provided a quantitative interpretability framework by measuring the agreement between the generated relevance maps and meta-learned disease likelihood maps, i.e., a proxy-ground truth ^23,24^. However, these were supervised approaches with only one fixed ground truth for all patients, i.e., the regional disease likelihood. Our study on the other hand, adopts an unsupervised approach that uses the morphological features as proxy ground truth features, which are unique to each patient. This allows for validation of the relevance maps based on the pathologic features tailored to each patient.

Based on the results presented in Table 2, we found that the inclusion of contextual information enhances the homogeneity of the clusters. Clustering in the enriched explanation space leads to better alignment with disease diagnosis labels. There is an improvement in the homogeneity (from 0.34 to 0.4) and V-measure (from 0.39 to 0.43), when comparing clustering outcome in enriched explanation space to basic explanation space. Our findings suggests that contextual features create relatively more coherent clusters, where now participants with the same disease diagnosis are clustered together. As a result, this refines the explanation space itself, making it more representative of the underlying disease pathology. However, the improved homogeneity comes at the cost of cluster separability, as shown by the lower silhouette score and increased DBI, suggesting a trade-off between interpretability and structural distinction in the explanation space.

To further assess the added value of CNN-derived features, we conducted an additional clustering experiment using only volumetric and cortical thickness measures as the feature space (see Table 2). The resulting clustering outcomes in this explanation space were subpar in terms of cluster homogeneity (0.26) and had limited ability to distinguish between disease stages (V-measure of 0.32). These findings suggest that while morphological features provide supportive contextual information by enriching the explanation space.

Clustering, unlike supervised overlap quantifications, also serves as a flexible approach for integrating diverse information sources, making it adaptable for future applications incorporating various pathological measures, e.g., by adding FDG-PET or tau-PET scans ^74,75^. This would allow for explanations to be generated from multi-modal data sources, possibly better capturing the interaction between various clinical factors and making the explanations more inclusive of diverse clinical contexts.

#### Group-based explanations

The identified subclusters in the context-enriched explanation space provided meaningful differentiation in longitudinal cognitive trajectories, reinforcing the importance of the CNN model’s attribution maps when grouped with morphological features. Participants in the stable subcluster demonstrated a significantly lower risk of progression, as evidenced by mixed-effects modeling (Figure 4) and Kaplan-Meier analysis (Figure 5), respectively. On the other hand, in the converter subcluster, participants were more likely to have a rapid cognitive decline. These findings highlight the potential of the clustering model to stratify participants’ disease progression risk, using structural MRI scans and CNN models trained on it, aiding in early identification and intervention planning. These explanations serve as simplified interpretations for generated relevance maps from CNN’s predictions and, without overly highlighting individual morphological or relevance features.

#### Explanation-by-examples

We used the K-nearest neighbor (KNN) model within the context-enriched explanation space to provide a dynamic method for generating example-based explanations of possible cognitive trajectories. Rather than relying on identified hierarchical sub-clusters, KNN allows for a dynamic selection of the neighborhood. By identifying participants with the most similar pathology, this method enables personalized projections of possible cognitive trajectories without making any modeling assumptions, as outlined by an earlier study ^24^.

The choice of KNN over alternative meta-models was intentional, as it abstracts away complex aggregation details that could obscure interpretability for a clinical user. For instance, explanations offered by interpreting a regression model’s parameters, i.e., beta coefficients may less intuitive and might not provide needed decision support functionalities. A similar argument for a lack of intuitive clarity could also be made about marginal contribution scores calculated via Shapley values. Instead, the chosen neighborhood-based approach offers a more accessible way to present likely disease trajectories by linking a participant’s current pathology profile to other participants tracked longitudinally. More importantly, the objective of the KNN model was not to develop a meta-classifier superior to the original CNN, but rather to offer example-based explanations that enhance interpretability. The notion of neighborhood plays a key role here, providing transparent and participant-specific insights into the rationale for predicting disease progression.

#### Rule-based textual explanations

Moving away from data-driven explanations towards knowledge-driven explanations, the ontology-based explanation method provides a structured approach to generating individualized textual summaries of neuroanatomical abnormalities. Rule-based summarization reduces cognitive overload on clinicians by hierarchically aggregating pathological findings using a-priori neuroanatomical knowledge. The generated template-based textual reports provide an intuitive means of communicating the model’s decisions to the clinical users. Unlike purely data-driven deep learning models, which often lack transparency, this approach integrates CNN relevance with morphological features in a rule-based manner, enhancing clinical usability.

Both rule-based explanations and explanation-by-examples generate so-called *local* explanations, which means they show individual properties of a single participant’s data. In contrast, methods that generate *global* explanations, which target the overall behavior of the whole model, might overlook the subtleties of individual cases. Local interpretations are often found to assist in making context-sensitive decisions ^76,77^, which is crucial in domains such as medical diagnosis.

### Qualitative evaluation of the explanation types

In our the focus-group interviews, we aimed to facilitate the collaboration between method developers and healthcare professionals. Martin et al.^11^ highlight the need for clinical stakeholders in evaluating XAI for dementia and radiology. Limited expert involvement hinders adoption and reduces the effectiveness of XAI methods, as clinicians ensure that the explanations align with their workflows and aid decision-making.

We report that the neurologists in our study favored group-level explanations for expert communication and risk assessment, but they were cautious about using explanation-by-example with patients. The radiologists in our sample preferred textual explanations for their workflow. Also, they viewed relevance heatmaps to be less useful for pathology reporting. These distinctions between the two professional groups underscores their differing priorities, with neurologists focusing on both current and future patient care, while radiologists concentrate more on the accurate description of pathological imaging findings to support diagnosis and treatment planning. Future XAI development in neurodegenerative research will benefit by accounting for these varying needs across clinical specialties and use cases.

### Limitations and future work

As the current work is based on a data-driven methodology, one key limitation is that the explanation space is inherently dependent on the CNN model and the relevance heatmap generation XAI method used for its creation. This implies that the subsequent quantification of explanations’ quality relies upon the performance of the underlying CNN model and relevance attribution method, which necessitates the use of a well-trained and generalizable CNN model to derive relevance attributions from. In the current study to mitigate this issue, during cross-validation we select the CNN model from the fold with the best performance metrics. Additionally, the cluster quality metrics within various explanation spaces (Table 2), are relative measures and should therefore be interpreted in relation to one another rather than as absolute indicators. Although, the qualitative evaluation of XAI methods highlighted key considerations for future research and development, there remains certain limitations. We acknowledge a small sample size of experts in our study. The selection of neurologists in was non-random, which may introduce selection bias, as those experts may already favor XAI adoption. These drawbacks would be rectified in our future work.

A limitation of our study is that mutual information based feature selection and downstream analysis used the same dataset. Although cross-validation with Fleiss’ Kappa showed stable outcomes, future work would be further strengthened by validation on independent datasets. Moving forward, future studies should also explore different XAI methods for relevance map generation and compare them head-to-head with the LRP method presented in our current study. More pertinently, our future research will focus on assessing the model’s and the generated explanation’s confidence and certainty, for we assume that this would enhance the reliability of the explanations ^43,49^. Furthermore, in future work we would like the explanations to be automatically tailored to their intended use case, i.e., distinguishing between communication among clinicians, where explanations are detailed, versus communication between clinicians and laypersons, which requires simplified and layperson-friendly language.

Another line of promising research is leveraging the large language models (LLMs) for knowledge-driven, ontology-based textual explanation refinement. Retrieval augmented generation (RAG) might be particularly suitable, as it improves interpretability by keeping LLMs grounded in the context provided^78^. This approach would minimize the risk of “hallucination” that is often associated with LLMs, while ensuring faithfulness to the underlying domain logic.

## Conclusion

This study introduces a framework for generating various types of explanations based on different XAI methods. Our proposed methods enrich the standard explanation space with clinically relevant morphological features. Our results demonstrate that the enriched explanation space yields more clinically meaningful insights, as shown by improved clustering metrics and the ability to distinguish between stable and converter participant subgroups. The explanation-by-example method visualizes exemplary possible cognition trajectories for a query participant for up to 72 months without making further modeling assumptions. The ontology-based textual explanations are dynamically generated in a rule-based manner, creating structured summaries that reduce cognitive overload for clinicians. Furthermore, our qualitative evaluation with clinicians highlighted the practical relevance of different explanation types.

## Author Contributions

Devesh Singh: conceptualization, investigation, methodology, software, visualization, writing – original draft; Stefan J. Teipel: writing – review & editing, supervision; Martin Dyrba: writing – review & editing, conceptualization, software, supervision; All other co-authors - Yusuf Brima, Fedor Levin, Martin Becker, Bjarne Hiller, Andreas Hermann, Irene Villar- Munoz, Lukas Beichert, Alexander Bernhardt, Katharina Buerger, Michaela Butryn, Peter Dechen, Emrah Düzel, Michael Ewers, Klaus Fliessbach, Silka D. Freiesleben, Wenzel Glanz, Stefan Hetzer, Daniel Janowitz, Doreen Görß, Ingo Kilimann, Okka Kimmich, Christoph Laske, Johannes Levin, Andrea Lohse, Falk Luesebrink, Matthias Munk, Robert Perneczky, Oliver Peters, Lukas Preis, Josef Priller, Johannes Prudlo, Diana Prychynenko, Boris S. Rauchmann, Ayda Rostamzadeh, Nina Roy-Kluth, Klaus Scheffler, Anja Schneider, Louise Droste, Björn H. Schott, Annika Spottke, Matthis Synofzik, Jens Wiltfang, Frank Jessen, Marc-André Weber: data acquisition, collection and curation, writing – review & editing.

## Acknowledgments

We sincerely appreciate the expertise and support of Dr. Großmann, Dr. Jäschke, Dr. Beller and Dr. Streckenbach from the Institute for Diagnostic and Interventional Radiology, Pediatric and Neuroradiology at the University Hospital of Rostock in the qualitative evaluation of the methods developed.

The data samples were partly collected from the DELCODE study group of the Clinical Research Unit of the German Center for Neurodegenerative Diseases (DZNE). Details and participating sites can be found at www.dzne.de/en/research/studies/clinical-studies/delcode. The data samples were partly collected from the DESCRIBE-FTD study group of the Clinical Research Unit of the German Center for Neurodegenerative Diseases (DZNE). Details and participating sites can be found at https://www.dzne.de/en/research/studies/clinical-studies/describe-ftd/. Data collection and sharing for this project was partially funded by the Alzheimer’s Disease Neuroimaging Initiative (ADNI) (National Institutes of Health Grant U01 AG024904). ADNI is funded by the National Institute on Aging, the National Institute of Biomedical Imaging and Bioengineering and generous support of other industry partners. A complete listing of ADNI investigators can be found at https://adni.loni.usc.edu/wp-content/uploads/how_to_apply/ADNI_Acknowledgement_List.pdf. Data was collected by the AIBL study group. AIBL researchers are listed at https://aibl.csiro.au. European DTI study on dementia EDSD was collected by nine European centers: Amsterdam (Netherlands), Brescia (Italy), Dublin (Ireland), Frankfurt (Germany), Freiburg (Germany), Milano (Italy), Mainz (Germany), Munich (Germany), and Rostock (Germany). FTLDNI was funded through the National Institute of Aging with the goal of identifying neuroimaging modalities and methods for tracking frontotemporal lobar degeneration (FTLD). For up-to-date information on participation and protocol, please visit http://memory.ucsf.edu/research/studies/nifd. Data collection and sharing for this project was funded by the Frontotemporal Lobar Degeneration Neuroimaging Initiative (National Institutes of Health Grant R01 AG032306).

## Data Availability Statement

The source code is available via GitHub: https://github.com/martindyrba/xai4dementia-framework

Data used for training/evaluation of the models is available from the respective initiatives ADNI: https://adni.loni.usc.edu/data-samples/, AIBL: https://aibl.org.au/, DELCODE: https://www.dzne.de/en/research/studies/clinical-studies/delcode, DESCRIBE: https://www.dzne.de/en/research/studies/clinical-studies/describe/, EDSD: https://www.gaaindata.org/partner/EDSD, NIFD/FTLDNI: https://memory.ucsf.edu/research-trials/research/allftd.

## Ethics Statement

The studies involving humans were approved by the respective neuroimaging initiatives internal review boards of each of the participating study sites. See https://adni.loni.usc.edu and https://aibl.org.au for details. All initiatives met common ethical standards in the collection of the data such as the Declaration of Helsinki. Analysis of the data was approved by the internal review board of the Rostock University Medical Center, reference number A 2020-0182. The studies were conducted in accordance with the local legislation and institutional requirements. Written informed consent for participation was not required from the participants or the participants’ legal guardians/next of kin in accordance with the national legislation and institutional requirements.

The study involved focus-group interviews with clinical experts who are cognitively normal individuals and for whom oral consent was deemed sufficient. First, the group supervisors from the Institute of Diagnostic and Interventional Radiology, Pediatric Radiology and Neuroradiology (IfDIR) and the Klinik für Psychosomatische Medizin und Psychotherapie (KPM-Geron) of the University Medical Centre Rostock were contacted with the interview request, explaining the study’s purpose and procedure. Individual experts were contacted for scheduling appointments. Before the start of the interviews, experts provided consent for both the interview and audio recording.

## Supplementary Material

### A.1 Neuroimaging datasets

**Supplementary Table A.1.1.** Patient statistics separated by the diagnosis group. The statistics are reported for each of the seven data cohorts. The patients were pooled from the following study cohorts: ADNI phase 2 and phase 3, AIBL, DELCODE, DESCRIBE, EDSD, and NIFD. CN: cognitively normal, MCI: mild cognitive impairment, AD: dementia due to Alzheimer’s disease, FTD: Frontotemporal dementia, where phenotypes include, BV: behavioral variant of FTD, SV: semantic variant of FTD, and PFNA: progressive nonfluent aphasia. MMSE: mini-mental state examination score, F: female, M: male. Numbers are reported as (mean ± sd).

|  |  |  |  |  |
| --- | --- | --- | --- | --- |
| **ADNI** | **CN** | **MCI** | **AD** | **FTD** |
| **Age** | 72.48 ± 6.99 | 73.20 ± 7.92 | 74.94 ± 7.90 |  |
| **MMSE** | 29.11 ± 1.13 | 27.70 ± 1.93 | 22.68 ± 3.25 |  |
| **Sex (M/F)** | 239/341 | 224/174 | 142/107 |  |
| **AIBL** |  |  |  |  |
| **Age** | 72.44 ± 6.18 | 74.33 ± 6.85 | 73.18 ± 7.26 |  |
| **MMSE** | 28.73 ± 1.23 | 27.04 ± 2.19 | 21.24 ± 5.31 |  |
| **Sex (M/F)** | 188/260 | 49/47 | 26/36 |  |
| **DELCODE** |  |  |  |  |
| **Age** | 68.25 ± 5.43 | 72.46 ± 5.72 | 74.66 ± 6.24 |  |
| **MMSE** | 29.47 ± 0.83 | 27.80 ± 1.97 | 23.09 ± 3.25 |  |
| **Sex (M/F)** | 120/162 | 81/72 | 42/61 |  |
| **EDSD** |  |  |  |  |
| **Age** | 68.59 ± 6.00 | 71.11 ± 7.41 | 72.69 ± 8.29 |  |
| **MMSE** | 28.61 ± 3.18 | 26.34 ± 2.99 | 20.86 ± 5.28 |  |
| **Sex (M/F)** | 98/97 | 94/76 | 56/78 |  |
| **NIFD** |  |  |  |  |
| **Age** | 62.67 ± 7.25 |  |  | 63.01 ± 7.08 |
| **MMSE** | 29.38 ± 0.77 |  |  | 23.79 ± 7.21 |
| **Sex (M/F)** | 58/74 |  |  | 75/57 |
| **DESCRIBE** |  |  |  |  |
| **Age** | 60.57 ± 13.40 |  |  | 64.08 ± 9.50 |
| **MMSE** | 29.07 ± 1.15 |  |  | 23.04 ± 6.87 |
| **Sex (M/F)** | 29/29 |  |  | 37/24 |
| **NIFD** | **CN** | **BV** | **SV** | **PFNA** |
| **Age** | 62.67 ± 7.25 | 60.97 ± 6.59 | 62.65 ± 6.10 | 67.44 ± 7.29 |
| **MMSE** | 29.38 ± 0.77 | 23.87 ± 5.66 | 22.59 ± 9.57 | 25.03 ± 6.78 |
| **Sex (M/F)** | 58/74 | 42/21 | 21/16 | 12/20 |
| **DESCRIBE** | **CN** | **BV** | **SV** | **PFNA** |
| **Age** | 60.57 ± 13.40 | 64.08 ± 9.50 |  |  |
| **MMSE** | 29.07 ± 1.15 | 23.04 ± 6.87 |  |  |
| **Sex (M/F)** | 29/29 | 37/24 |  |  |

### A.2 Model Training

We trained a DenseNet model, using a stratified five-fold cross-validation (see Fig. A.2.1). The models were trained for a three-way classification - AD-vs-CN-vs-FTD. Here Alzheimer’s dementia (AD) patients and patients with amnestic mild cognitive impairment (MCI) were merged into one disease-positive class, while multiple phenotypes of frontotemporal dementia (FTD) - behavioral variant (bvFTD), semantic dementia (SD), and progressive nonfluent aphasia (PNFA) were also clubbed under one FTD class. These two classes were compared against the cognitively normal (CN) participants, i.e., the control class.

Categorical cross-entropy was chosen as the loss function. The models were optimized using the Adam optimizer with a learning rate of 0.0001, and other parameter settings were set to default. We trained the models for 100 epochs, using a batch size of 128. To reduce model over-fitting, an early stopping regularization method was applied, monitoring the validation set loss as a performance metric over epochs, with patience of 5 epochs and a minimum change threshold of 0.01. To avoid overfitting, we also weighted the model’s error with the label’s class weight. During each cross-validation run, only the best-performing model was saved.

For training, the data augmentations were generated using AUCMEDI Python package, where 3D volumes were randomly left/right-flipped with a 50% probability, and rescaled with a 50% probability within the zooming in or out limits of 90% and 110%. These augmentations were only applied during model training and were disabled on validation and test sets.

**Supplementary Figure A.2.1.**
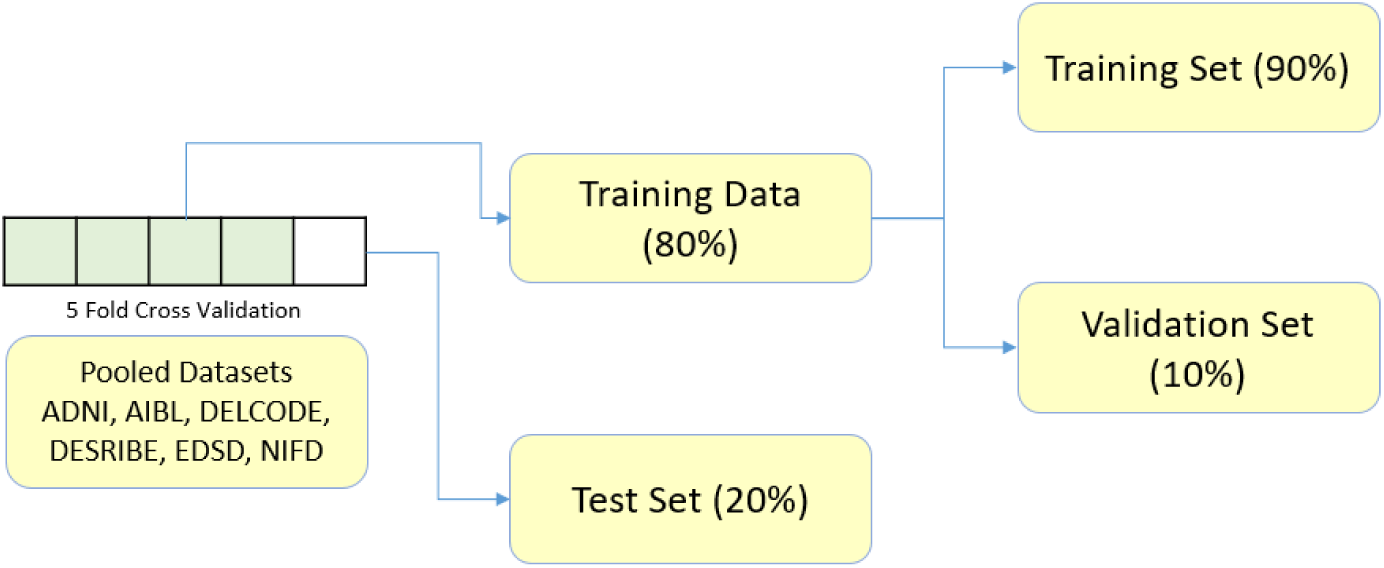
Schematics representation of the data splitting for CNN model training.

Based on the results from the 5-fold cross-validation training of the models (see Table A.2.1), we chose fold 1 as the default model for further analysis in our study. The model training results from fold 1 are illustrated in Figure A.2.2.

**Supplementary Table A.2.1.**
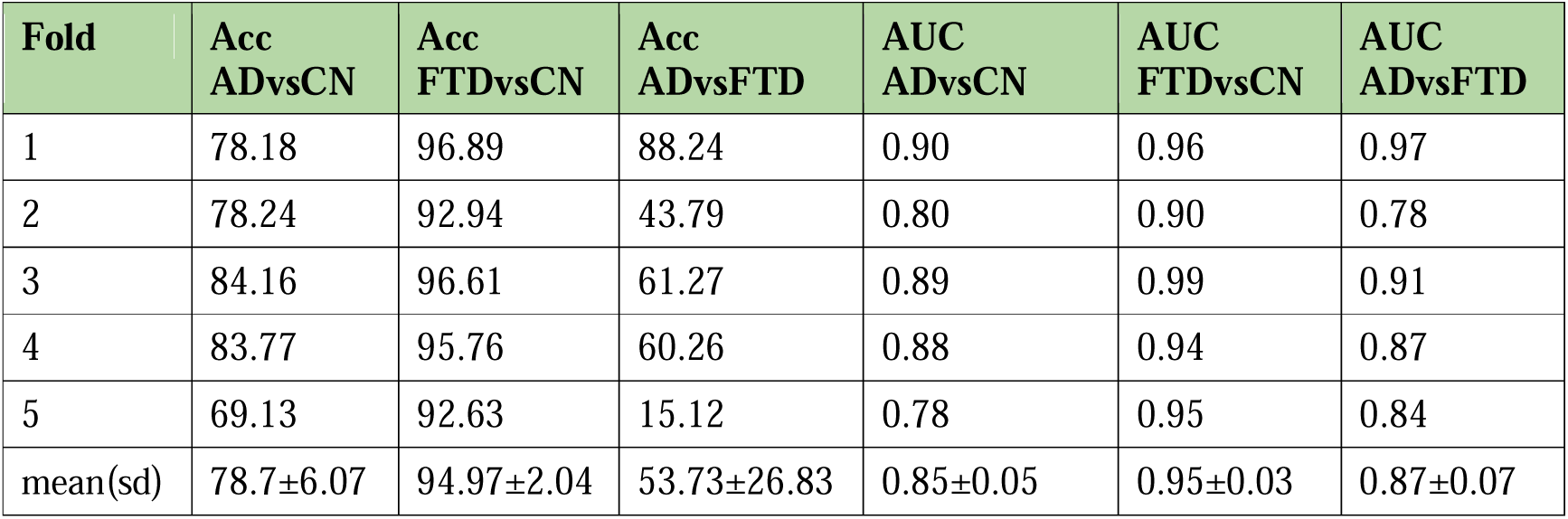
Performance metrics on the test set. Acc: simple accuracy, AUC: Area under the (ROC) curve. CN: cognitively normal, AD: dementia due to Alzheimer’s disease (which due to design choices, also includes the amnestic mild cognitive impairment (MCI) subjects), and FTD: frontotemporal dementia.

**Supplementary Figure A.2.2.**
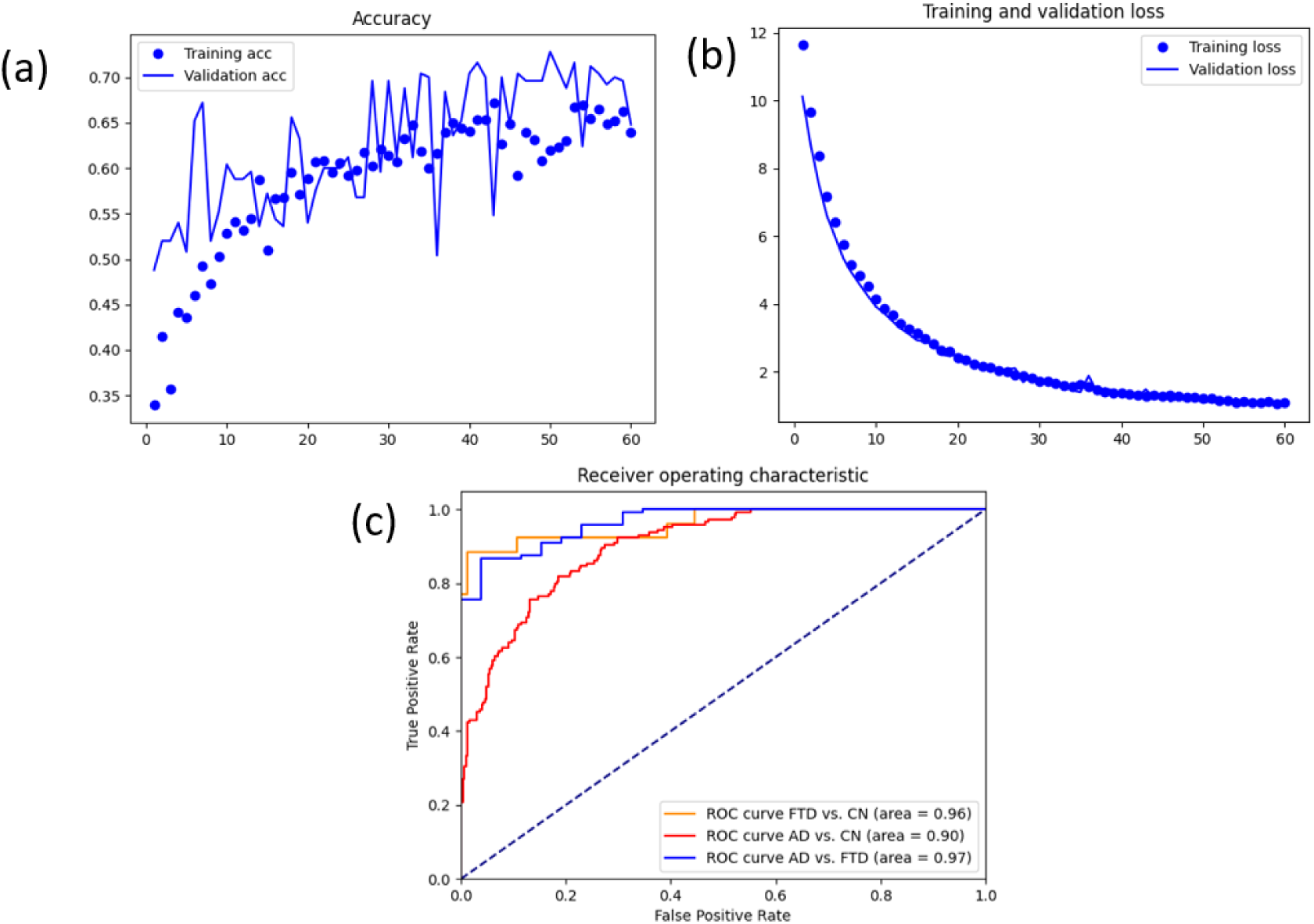
Model training results from fold 1: (a) (simple) Accuracy metric on train and validation set, (b) loss metric on train and validation set, and (c) binarized ROC-AUC curves on the test set. CN: cognitively normal, AD: dementia due to Alzheimer’s disease (which due to design choices, also includes the amnestic mild cognitive impairment (MCI) subjects), and FTD: frontotemporal dementia.

The mean relevance maps for the test set of fold 1 are visualized below. For relevance attribution, we employed the compositional LRP rule (= 1, β = 0), as established in our previous work for generating clinically meaningful explanations^1^. To enhance the signal-to-noise ratio during visualization, we re-scaled the relevance intensities based on the 99.99^th^ percentile (q = 0.9999) and clipped the resulting values to the range [−1, 1]. A Gaussian smoothing filter with a standard deviation of 0.8 was then applied to further improve interpretability.

The mean relevance maps of Alzheimer’s disease (AD) dementia and mild cognitive impairment (MCI) patients appeared visually similar; however, distinct patterns emerged when comparing across disease groups. In the AD group, supplementary figure A.2.3, relevance was concentrated in the hippocampus (slices [-20, -10]) and bilaterally in the thalamus (slices [-30, -20]). In contrast, the frontotemporal dementia (FTD) group, supplementary figure A.2.4, exhibited prominent relevance in the frontal lobes, particularly the right insula and frontal opercular cortex in slice -8, as well as the pregenual anterior cingulate cortex (pACC) in slices [37, 44]. Notably, insular involvement was also reported in our prior study^2^, suggesting consistency across different model training strategies and relevance attribution techniques in identifying clinically relevant brain regions.

**Supplementary Figure A.2.3:**
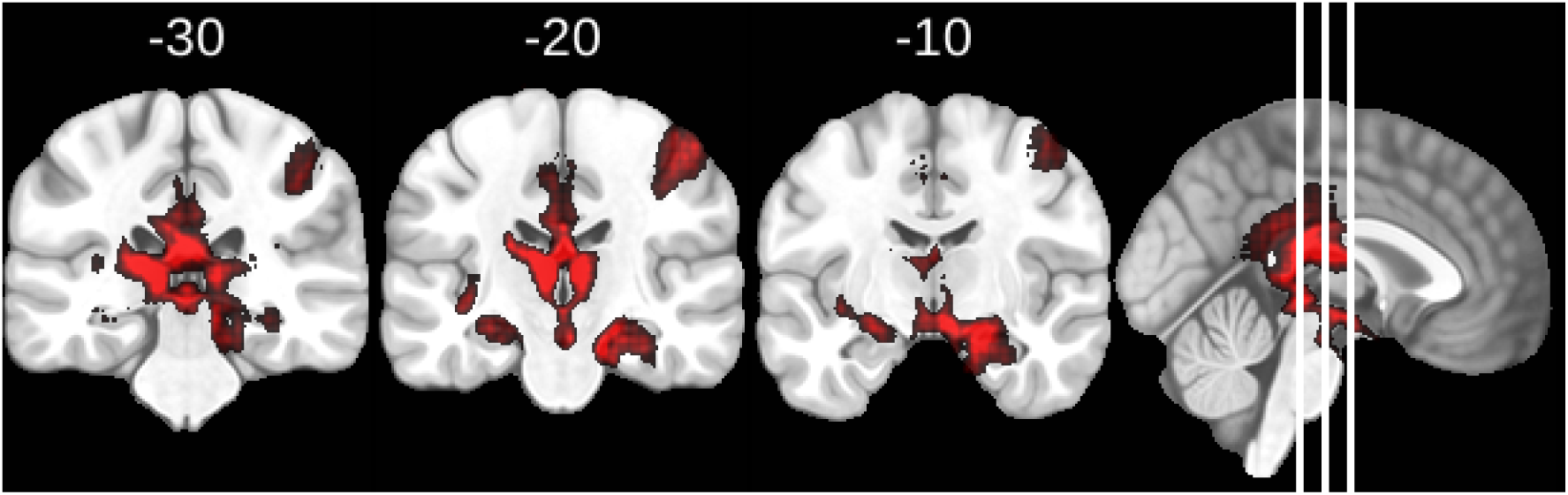
Mean relevance maps for the AD group of the test dataset obtained using the LRPa=1,β=0 relevance propagation method overlaid on MNI brain template. Coronal slices show Y=[-10,-20,-30] mm in MNI reference space are shown. The most relevant input regions are highlighted. Relevance maps were created following proportional scaling of the activations.

**Supplementary Figure A.2.4:**
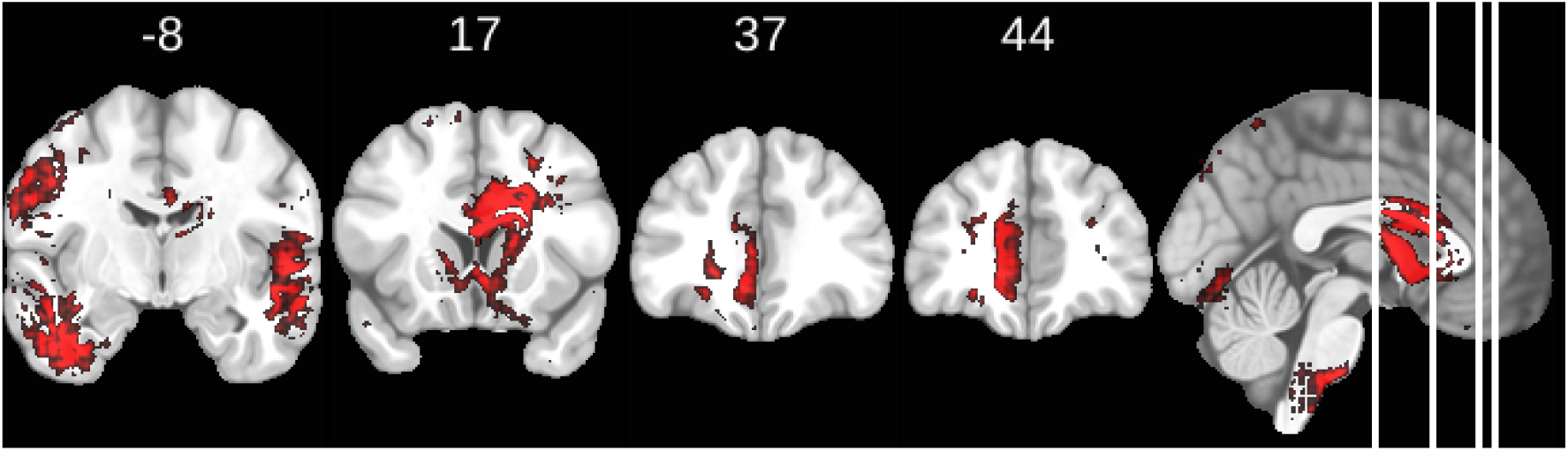
Mean relevance maps for the FTD group of the test dataset obtained using the LRPa=1,β=0 relevance propagation method overlaid on MNI brain template. Coronal slices show Y=[-8,17,37,44] mm in MNI reference space are shown. The most relevant input regions are highlighted. Relevance maps were created following proportional scaling of the activations.

## A.3 Feature Selection with Mutual Information

All features used in the mutual information analysis were derived from w-scores, representing age, sex, brain size and MRI scanner strength adjusted residualized values. Average cortical thickness measures were only estimated for cortical regions (e.g., superior temporal gyrus, frontal lobe areas) and not for subcortical structures (e.g., hippocampus, thalamus, or basal ganglia), which explains the absence of cortical thickness values in these regions.

To enhance transparency and accessibility, we have uploaded the intermediate results from our analysis pipeline to GitHub: https://github.com/martindyrba/xai4dementia-framework/. The repository includes a CSV file that specifies, for each of the 120 regions, which of the three feature types (CNN relevance, volumetry, or cortical thickness) passed the mutual information threshold and were included in downstream analysis. This is process outcome is also visualized in Supplementary Figure A.3.1.

**Supplementary Figure A.3.1:**
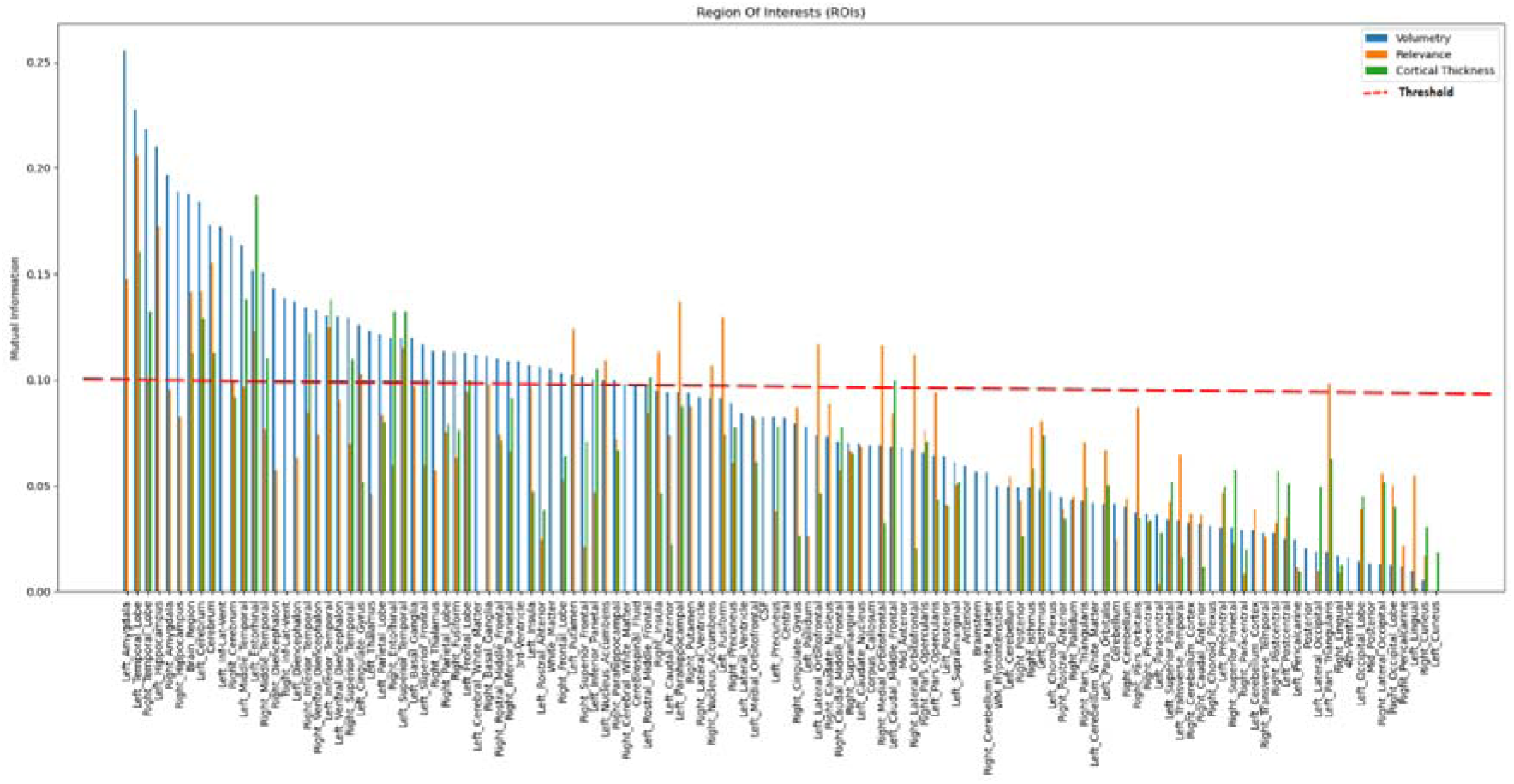
Mutual Information-Based Feature Selection: This figure illustrates the mutual information values computed across three different *w-score* features - CNN relevance, volumetry, and cortical thickness w-scores, representing their shared information content in comparison to the disease diagnosis label. The w-score features were sorted according to their mutual information on the volumetry features. The w-score features with mutual information above the threshold of 0.1 were retained as relevant and were selected for further analysis. For a vector graphic rendering, please refer to the GitHub version of the plot.

## A.4 Mixed-Effects Models of Cognitive Trajectories

Mixed-effects model experiments were done to investigate cognitive decline in patients, while accounting for repeated measures and inter-individual variability. The analysis was conducted using data from two groups: the Alzheimer’s Disease Neuroimaging Initiative (ADNI) and the DZNE Longitudinal Study on Cognitive Impairment and Dementia (DELCODE) cohorts. These datasets contain repeated cognitive assessments for each patient for up to 6 years, allowing for a longitudinal investigation of cognitive decline.

We tested a series of increasingly complex mixed-effects models. By incrementally adding predictors and interaction terms, we assessed model fit and explanatory power. The best-fitting model was determined using likelihood ratio tests via ANOVA.

**Model 1 (Base Model):** Includes age, sex, and the interaction between cluster membership and follow-up months (FUMonths) while accounting for repeated measures per participant.

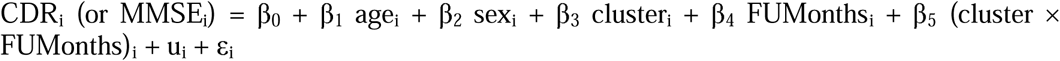

where u ∼ N(0, cr^2^) represents the random intercept for each participant, and E ∼ N(0, cr^2^) is the residual error term.

**Model 2 (Expanded Diagnosis Model):** Adds baseline diagnosis as a fixed effect.

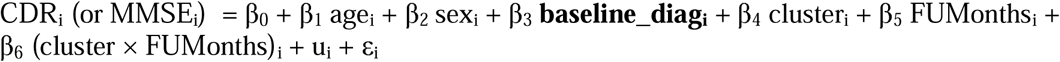

**Model 3 (Final Model):** Introduces an interaction between baseline diagnosis and follow-up months.

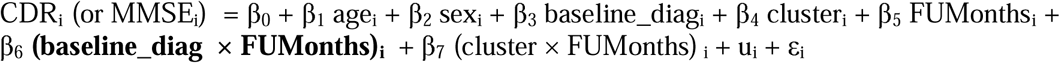

To compare model fit, ANOVA tests were performed, evaluating the nested model comparisons. The results indicated that Model 3 provided the best fit, suggesting that the interaction between baseline diagnosis and follow-up time significantly improves the model’s explanatory power. Specifically, for the CDR global (Figure A.4.1:), the high-risk converter group showed an annual increase of 0.074 points, while the low-risk group remained relatively stable with an increase of 0.007 points per year.

**Supplementary Figure A.4.1:**
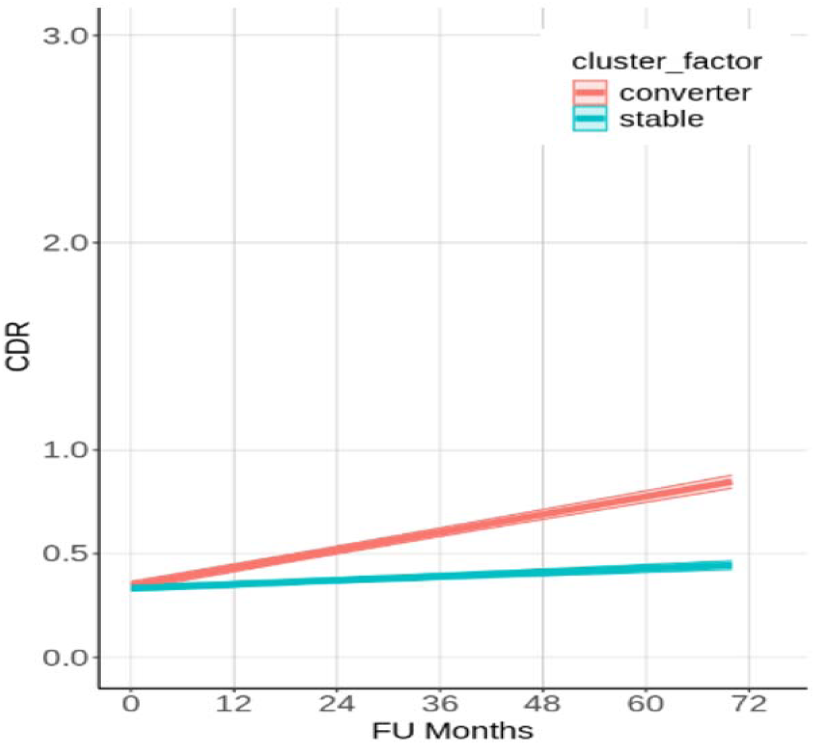
Longitudinal cognitive trajectories of different clusters of patients. Values on Clinical Dementia Rating (CDR) global are obtained from mixed effects regression models which included the age, sex, baseline disease diagnosis, and the interaction between cluster membership and follow-up time in months (FU Months), as well as the interaction between baseline disease diagnosis and follow-up months. The model also included random intercepts for each patient to account for repeated measurements. The shaded regions represent 95% confidence intervals.

## A.5 Explanation-by-example plots for Clinical Dementia Rating (CDR)

**Supplementary Figure A.5.1:**
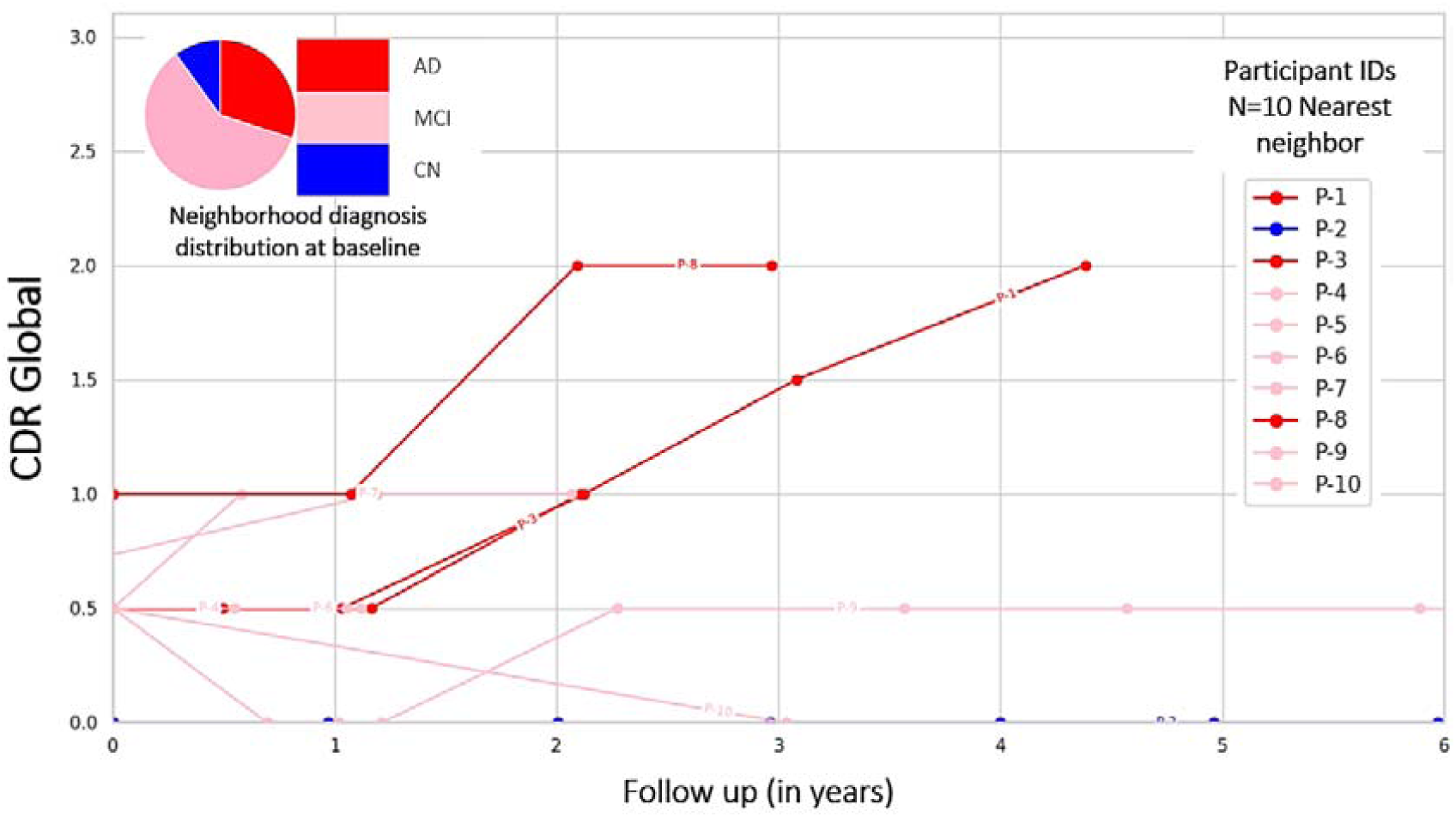
Explanation-by-examples: Within the context-enriched explanation space, the longitudinal cognitive trajectories of k=10 nearest neighbors of a query patient, from the DELCODE cohort, are shown. Patient IDs of the nearest neighbors are pseudonymised, and the nearest neighbors are listed in the order of increasing Euclidian distance from the query sample. Scores on the cognitive test Clinical Dementia Rating (CDR) global were observed on follow-up examinations for up to 6 years. The cognition trajectories are additionally color-coded by the baseline disease diagnosis.

## A.6 Explanation-by-example plots, each participant colored individually

**Supplementary Figure A.6.1:**
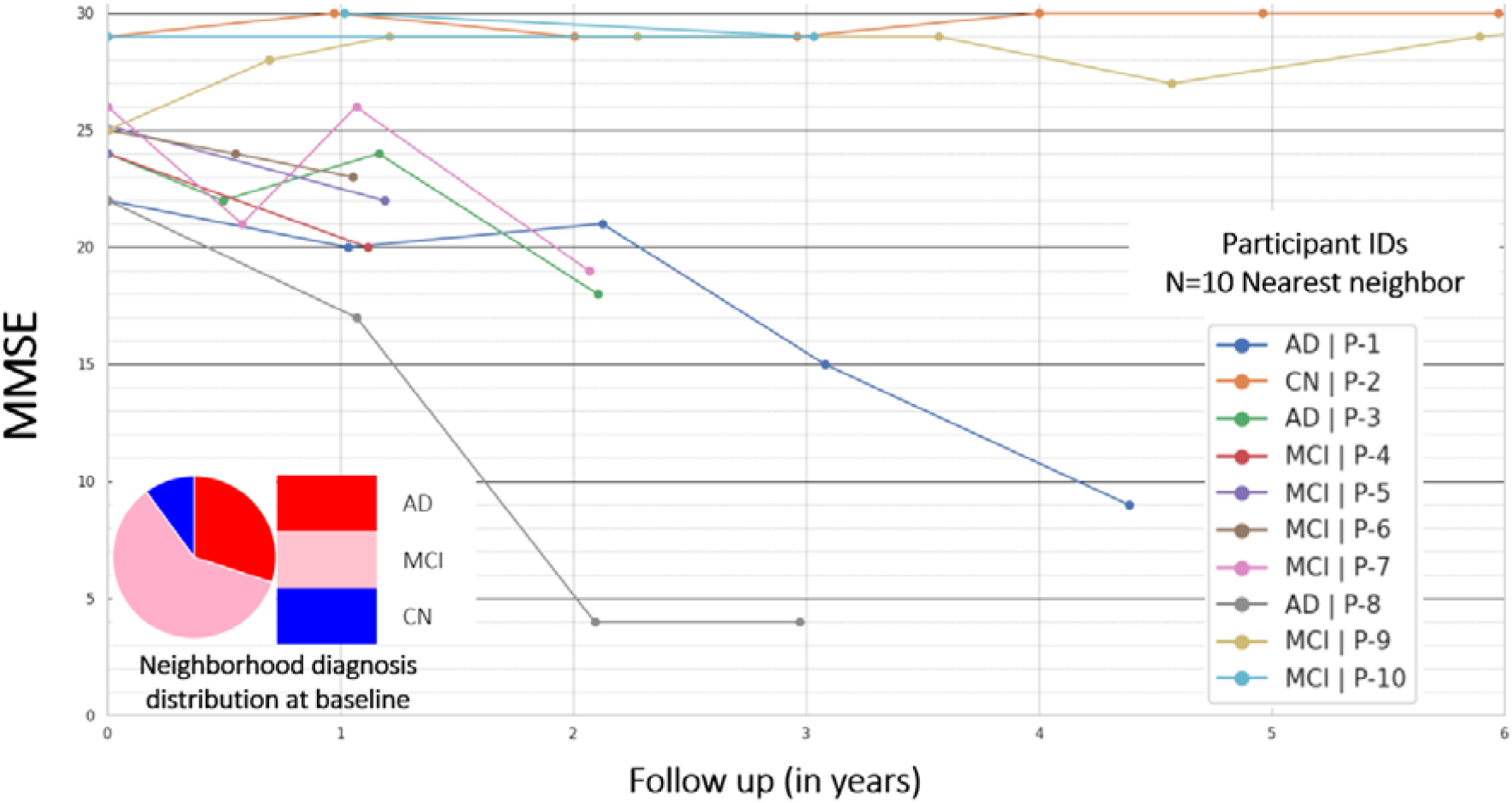
Explanation-by-examples: Longitudinal MMSE trajectories of the *k = 10* nearest neighbors of a query participant from the DELCODE cohort, observed for up to 6 years. Here, each cognitive trajectory is shown in a unique color for more detail.

**Supplementary Figure A.6.2:**
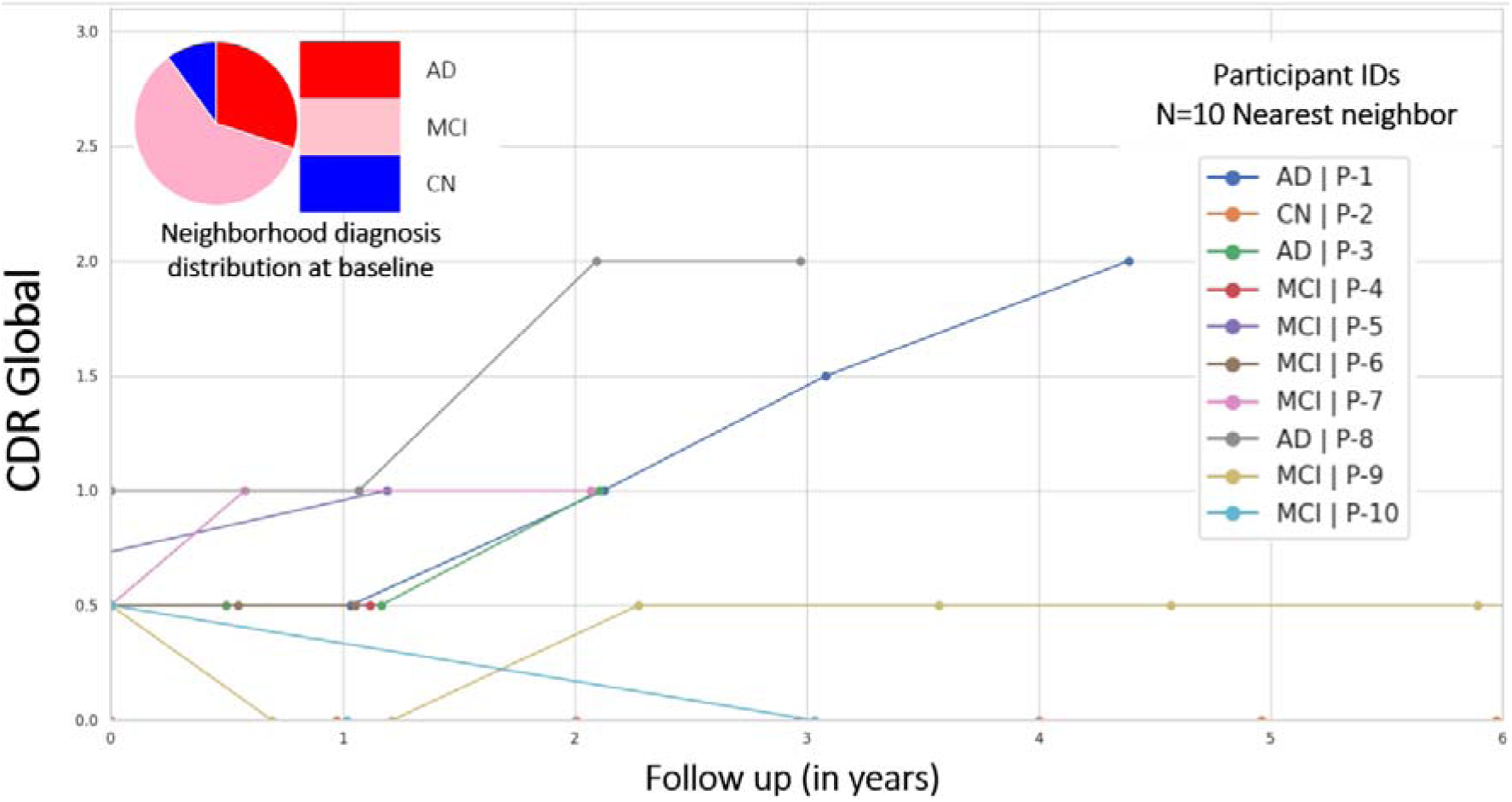
Explanation-by-examples: Longitudinal CDR trajectories of the *k = 10* nearest neighbors of a query participant from the DELCODE cohort, observed for up to 6 years. Here, each cognitive trajectory is shown in a unique color for more detail.

